# Comprehensive repertoire of the chromosomal alteration and mutational signatures across 16 cancer types from 10,983 cancer patients

**DOI:** 10.1101/2023.06.07.23290970

**Authors:** Andrew Everall, Avraam Tapinos, Aliah Hawari, Alex Cornish, Amit Sud, Daniel Chubb, Ben Kinnersley, Anna Frangou, Miguel Barquin, Josephine Jung, David N Church, Genomics England Research Consortium, Ludmil Alexandrov, Richard Houlston, Andreas J. Gruber, David C. Wedge

## Abstract

Whole genome sequencing (WGS) allows exploration of the complete compendium of oncogenic processes generating characteristic patterns of mutations. Mutational signatures provide clues to tumour aetiology and highlight potentially targetable pathway defects. Here, alongside single base substitution (SBS), double base substitution (DBS), small insertions and deletions (ID) and copy number aberration (CN) signatures covered by the Catalogue of Somatic Mutations in Cancer (COSMIC) we report signatures from an additional mutation type, structural variations (SV), all extracted *de-novo* from WGS in 10,983 patients across 16 tumour types recruited to the 100,000 genomes project. Across the five mutation classes we report 137 signatures, with 29 signatures new to COSMIC, including the first COSMIC SV signature reference set. We relate the signatures to clinical outcomes and likely response to therapy, demonstrating the role of signature analysis in delivering the vision of precision oncology.

## INTRODUCTION

Precision oncology aims to tailor therapy to the unique biology of a patient’s cancer to optimise treatment efficacy. Underpinning precision oncology is the concept of somatic mutations as the foundation of cancer development and the number of approved therapies for specific “actionable” mutations is increasing^1^.

Currently, multiple standalone tests or a panel are typically used to capture a set of genomic features for a given tumour type to guide precision oncology. However, falling costs make whole genome sequencing (WGS) a potentially attractive proposition as a single all-encompassing test. This approach is being explored by the 100,000 Genomes Project (100kGP), which is seeking to deliver the vision of precision oncology to National Health Service (NHS) patients as part of routine care^2^.

As well as identifying causative driver mutations, WGS allows exploration of the full landscape of mutations that describe the oncogenic processes, resulting in patterns – mutational signatures – which provide clues to tumour aetiology^3–6^. Moreover, mutational signatures are increasingly utilised to highlight potentially targetable pathway defects beyond reliance solely on driver mutations^7–10^. To deconvolute mutational signatures in cancer, mathematical methods estimate the number of mutations attributable to each signature. The Catalogue of Somatic Mutations in Cancer (COSMIC) provides an ongoing compendium of mutational signatures and, where this is known, their specific aetiology^3, 11–14^.

Here, we report mutational and chromosomal signatures extracted from WGS in 10,983 patients across 16 tumour types recruited to 100kGP. Across mutation classes we report 137 signatures, 29 of which are currently not catalogued by COSMIC. We relate new and known signatures to each other as well as to tumour characteristics and clinical outcomes to inform our understanding of oncogenesis, tumour progression and precision oncology.

## RESULTS

### The 100kGP cohort

The analysed cohort (100kGP, release v11) comprises tumour-normal sample pairs recruited to 100kGP through 13 NHS Genomic Medicine Centres across England. We restricted our analysis to samples with high-quality data from PCR-free fresh-frozen material, and samples that could be unambiguously assigned to a specific tumour histology. This yielded 10,983 samples from 10,975 participants (41 tumour histologies, across 16 tissue types). In addition to using variant calls from the 100kGP analysis pipeline we: (i) removed alignment bias introduced by ISAAC soft clipping of semi-aligned reads ^17^; (ii) called tumour copy number using Battenberg^18^; (iii) called SVs from the consensus of Manta^19^, LUMPY^20^, and DELLY^20, 21^; (iv) removed indels within 10 base pairs (bp) of a common germline indel. The final dataset comprised 285,233,025 somatic single base substitution (SBS) mutations, 2,272,616 doublet base substitution (DBS) mutations, 82,161,895 small insertions and deletions (ID), 1,285,875 copy number (CN) alterations and 811,962 structural variants (SVs). Information on sample-level and variant-level quality control are provided in the Methods and in **Supp. Table 1**.

### Mutational signature analysis

We extracted SBS, DBS, ID, CN and SV signatures independently for each mutation type and each of the 16 tissue types using SigProfilerExtractor (SPE)^22^. We extended the classification of SBS signatures from the conventional 96 classes to 288 by considering the transcriptional context of mutations; whether mutations fell on the transcribed, untranscribed or non-transcribed strand^22^. DBS, ID and CN mutational types were classified as *per* COSMIC. SV signatures are not currently included in the COSMIC database and were classified based on type and size of the SV and whether it was part of a cluster^15^. Pancancer, we extracted 67 SBS, 19 DBS, 18 ID, 20 CN and 13 SV signatures with signature activities shown in **Fig. 1**. These include 3 SBS, 8 DBS, 4 ID and 1 CN signatures not previously catalogued by COSMIC (**Fig. 2**). The 13 SV signatures were recovered by merging similar SV signatures discovered *de-novo* across multiple tumour types or using the *de-novo* signature where there are no similar signatures in other tumour types (**Methods P7**).

**Figure 1.**
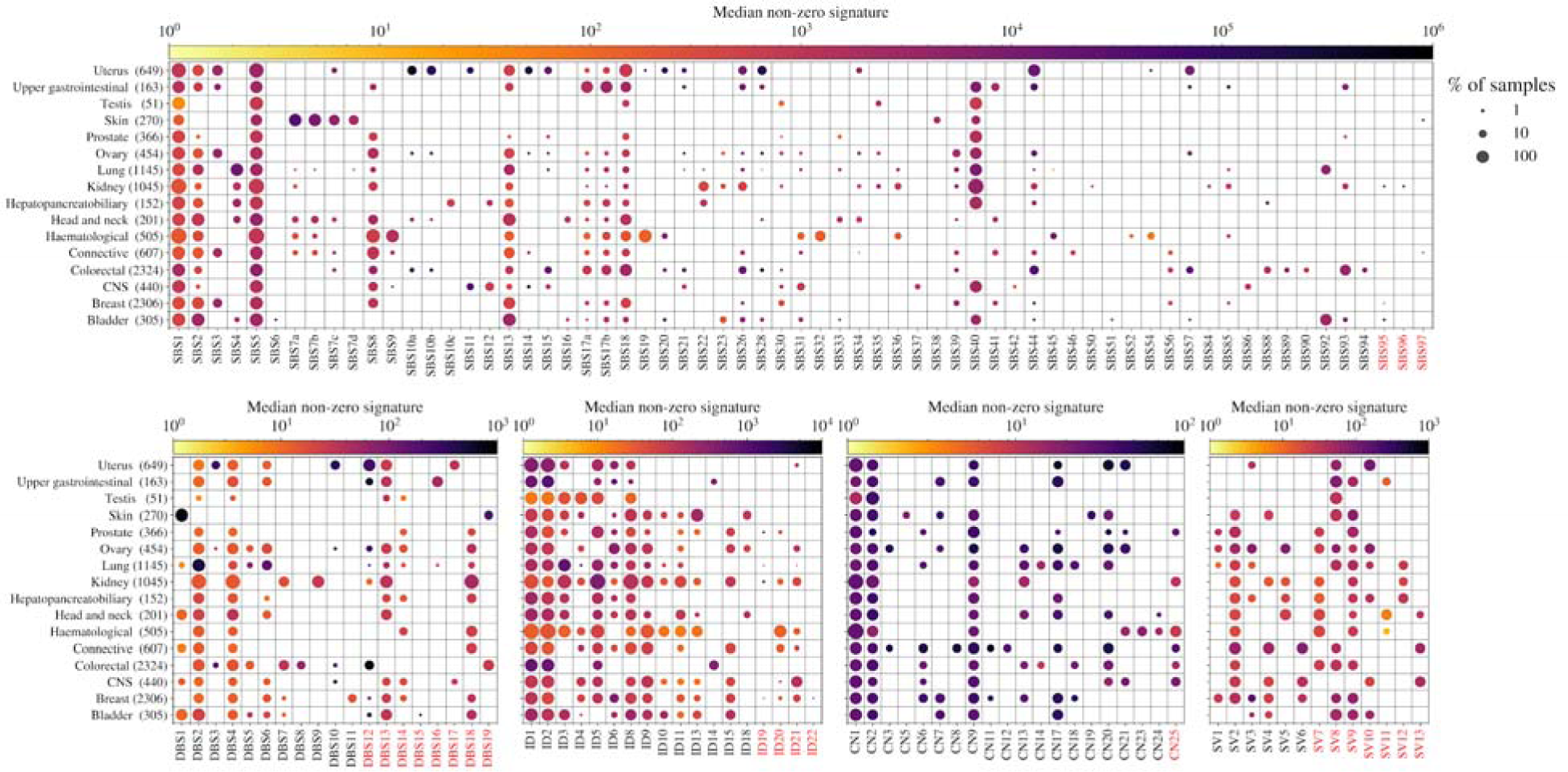
Signature activity across cancer types. Circle size corresponds to the percentage of samples in which the signature is non-zero while its colour corresponds to the median activity, i.e. mutation/alteration burden, of non-zero activity samples. Red labels highlight signatures not in COSMIC or not reported by Nik-Zainal *et al^5^*in the case of SV signatures. For each signature type, only samples with > 20 mutations/alterations are included to reduce noise.

**Figure 2.**
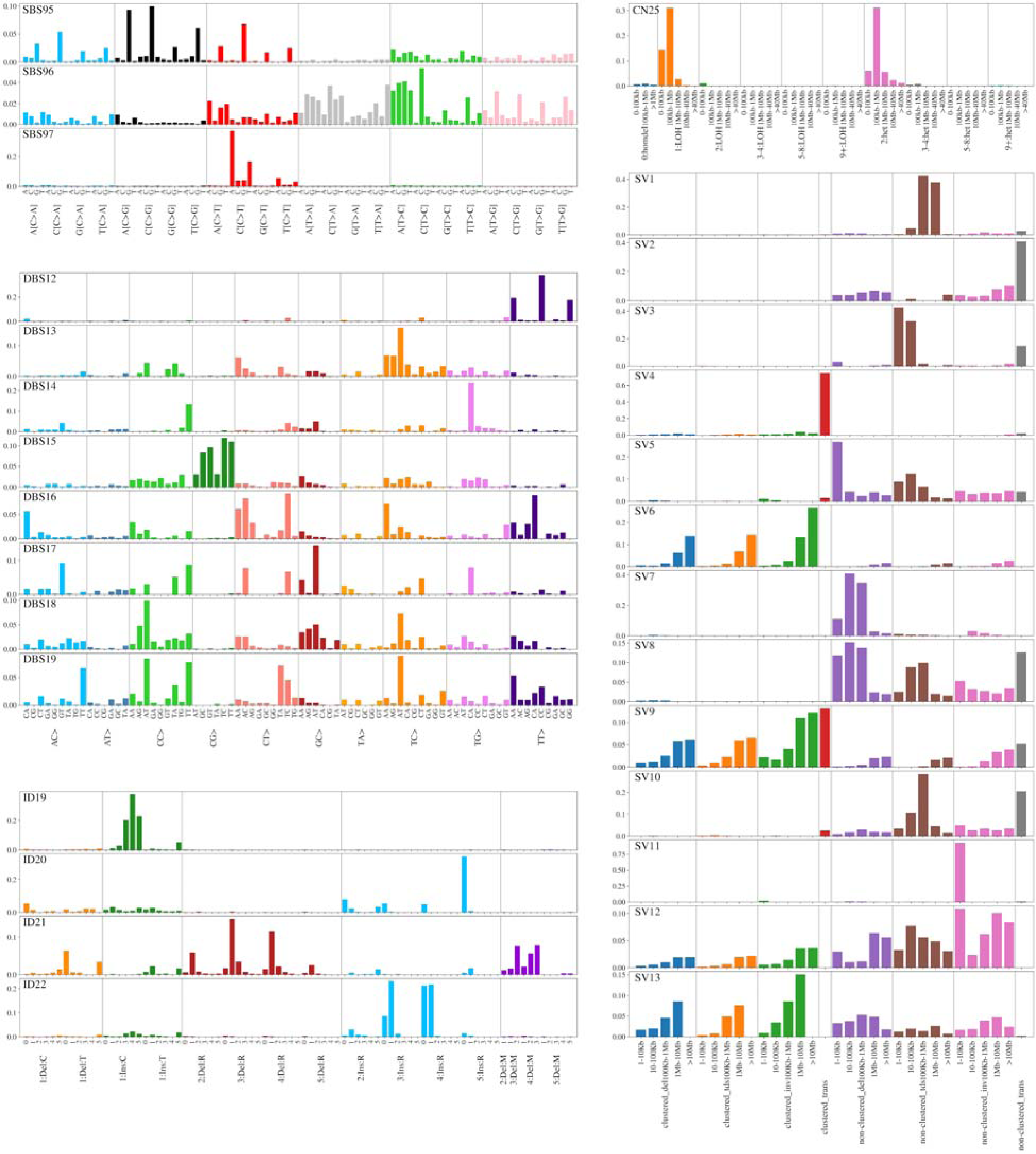
Novel extracted SBS, DBS, ID and CN signatures and 13 SV signatures extracted *de novo*. SV1-6 are matched to the 6 signatures reported by Nik-Zainal *et al^5^*.

### Characteristics of novel signatures

Of the 13 SV signatures, six are similar to those reported in a previous breast cancer study^5^ (cos(sim)>0.8) and we numbered these to match the previous study. In addition to breast cancer, SV1 and SV3 are also prevalent in ovarian cancers. SV4 and SV6 are a feature of connective tissue cancers and SV2 is present across multiple tumour types. Although SV5 was first identified in the breast cancer study we do not recover this signature in this cancer type in our analysis. SV7, SV8, SV11 and SV13 were previously reported in Degasperi *et al.*^15^. SV9 can be approximated by a linear combination of SV4, SV6 and SV13, however, it is still included in the reference list as it is independently extracted in 4 cohorts which have no SV4, SV6 or SV13 activity (**Supp. Materials P3**). Similarly, SV10 can also be approximately reconstructed from SV1 and SV2. SV12, which is novel herein, is primarily composed of both short and long inversions and is found in lung, kidney, mesothelioma, pancreas and bile duct cancers.

SBS95, seen in bladder, breast and kidney cancers, is dominated by mutations at an N<u>C</u>G context (**Fig. 2**). The SBS95 mutation profile shows similarity to SBS87, which in acute lymphoblastic leukaemia (ALL) has been linked to thiopurine treatment, albeit with C>T dominance^23^. SBS96, characterised by a broad mutation profile, is detected only in a minority of kidney cancers. SBS97, which is dominated by C>T mutations and shows similarity to SBS7b, is a feature of skin and connective tissue cancers. While new to COSMIC, these three SBS signatures have also been reported by Degasperi *et al*^16^.

DBS12 is an artifactual signature, caused by T>C mutations where in repetitive T regions, a flanking C is displaced due to read misalignment (reported by Degasperi *et al* ^16^ as DBS28; personal communication, see **Supp. Materials P2**). DBS13, composed primarily of mutations to TC dinucleotides, and associated with homologous recombination deficiency (HRD) signatures SBS3, ID6, CN17 and SV3, is identified in breast, ovarian and endometrial cancers. The CC>TT dinucleotide mutation, which characterises UV exposure^24^, is also a feature of DBS14 but is not extracted in melanoma. DBS15, which is characterised by mutations of CG dinucleotides, is seen in a subset of bladder cancer. DBS16 is composed of CA>AC and TA>AT mutations suggesting short inversion as its functional basis. DBS17 is seen in central nervous system (CNS) and uterine cancers and consists of a variety of mutations which all involve substituting GC for AT base pairs or vice versa. The primary mutations contributing to DBS18, NC>AT, are also the basis of SBS8 and SBS22, suggesting a common aetiology. DBS19 also includes C>T mutations, often paired with a T>A, which is the dominant contribution to SBS7c. DBS19, like SBS7c, is observed in skin and colorectal cancers (CRCs) suggesting that it can be produced either by UV exposure or another as yet unidentified mechanism. Of these 8 DBS signatures, DBS12-17 have cos(sim)>0.8 with signatures extracted by Degasperi *et al.*^16^ and DBS18 can be decomposed to other novel signatures (**Supp. Table 3**). DBS19 remains novel compared to previous studies and cannot be constructed from any other signatures.

Three of the four novel ID signatures, ID19, ID20 and ID22, involve base pair insertions - single base C insertion, 5bp insertion, and a 3 or 4 bp insertion. While ID19 and ID22 are both rare, ID20 is commonly extracted across haematological tumours. ID21 is unique in being composed of 2-4bp deletions in double-repeats clustered with ID9, SV4, SV6 and SV13, suggesting chromothripsis as a common basis (**Fig. 3**).

**Figure 3.**
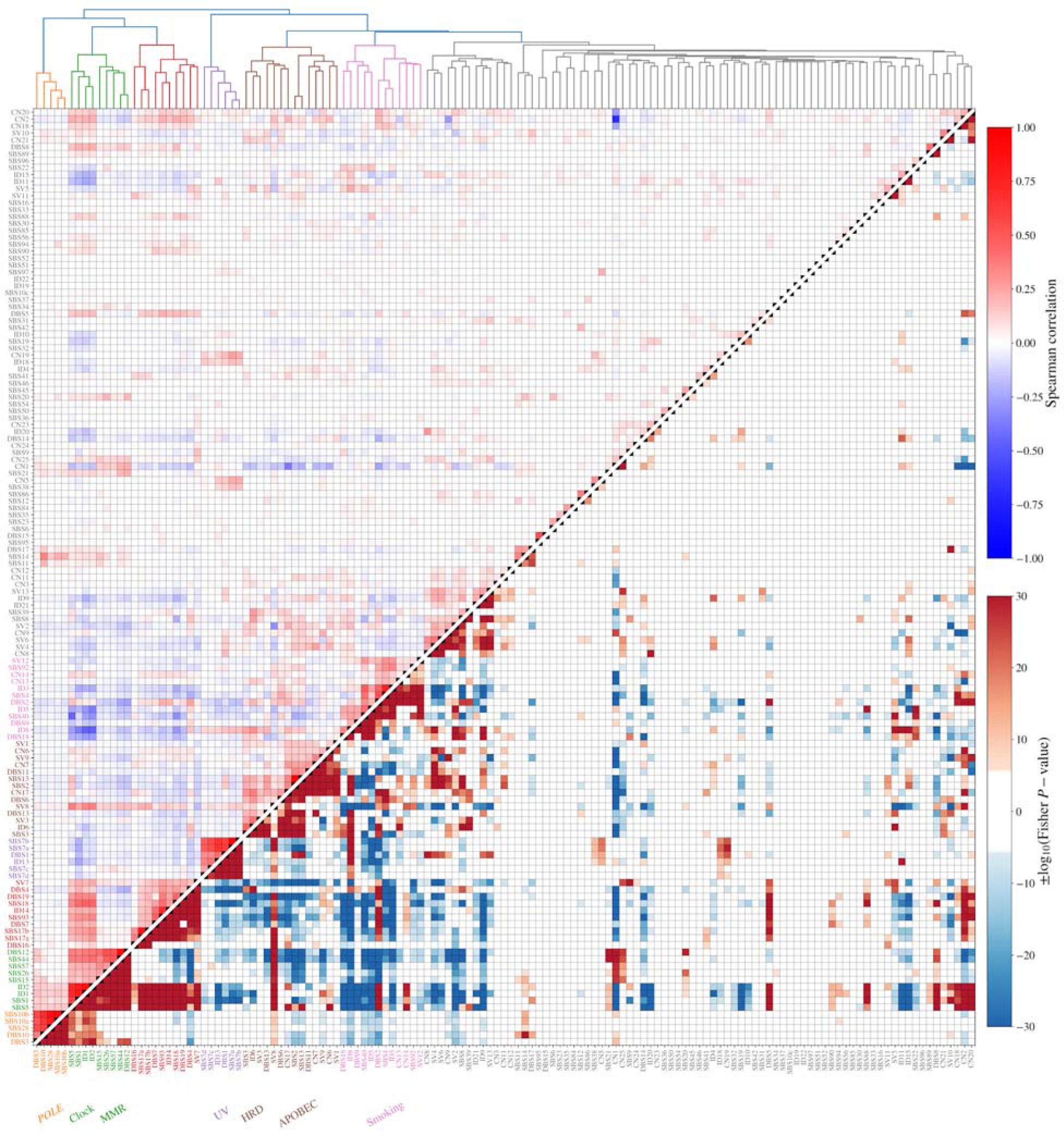
Relationship between SBS, DBS, ID, CN and SV signatures across cancers. Hierarchical clustering (Ward variance minimisation with Euclidean distance implemented in scipy) of all extracted signatures using log(activity + 1). Distinctly extracted clusters include UV, smoking, HRD, POLE and MMR. The upper triangle in the figure shows the Spearman correlation between log(activity + 1) for signatures across all samples with red squares showing correlated signatures. The lower triangle shows the log *P*-value of a two-sided Fisher exact test on the signature having non-zero activity (or, if more than half of the samples have non-zero activity, on the upper 50th percentile). Only those with Bonferroni-adjusted p-value<0.05 are shown and blue represents negative and red positive associations.

The novel CN signature, CN25, is a feature of CNS, connective tissue and prostate cancers and is associated with *MSH6* inactivation (**Fig. 5**). CN25 is composed of <1Mb loss of heterozygosity (LOH) deletions (similar to CN9 involving LoH >1Mb), which is indicative of chromothripsis. However, unlike other chromothripsis-associated CN signatures, CN4 - 8, CN25 is composed of low total copy number states with single copy LOH and diploid heterozygous states.

### Signature relationships

To further our understanding of tumorigenic processes we examined the interrelationship between signatures and their association with molecular and pathological phenotypes.

We observed signature clusters relating to UV exposure, smoking, APOBEC activity, deficient DNA mismatch repair (dMMR), HRD and polymerase epsilon (*POLE*) mutation^3, 4, 13, 25–27^, as well as clock-like signatures (**Fig. 3**). We identified a novel cluster of signatures including SBS17a, SBS17b, SBS18, SBS93, DBS4, DBS7, DBS16, DBS19, ID14 and SV7, which are active in many CRC and upper gastrointestinal samples. SBS1, SBS5 and ID2 are observed in the same samples as both dMMR and POLE signatures and the novel CRC cluster. For a number of signatures we found new inter-relationships suggesting a common aetiological basis. ID5, CN13, CN14 and SV12 are associated with smoking signatures SBS4, DBS2, ID3 and SBS92^28^. HRD and APOBEC signatures tended to co-occur as previously documented^29^ but are associated with the novel signature DBS13.

Within each tumour type some signatures are more common within a specific histology (**Fig. 4**). Ductal breast cancers are enriched for HRD signatures (SB3, ID6, CN17) as expected^30^ as well as ID8, which is indicative of defective non-homologous end joining (NHEJ). Chromophobe renal cancers (ChRCC) are enriched for HRD signature ID6, APOBEC SBS signatures (SBS2, SBS13) and the novel signature ID21, whereas papillary renal cancers are enriched for SBS22, which is caused by aristolochic acid exposure^6^. Signatures associated with the sarcomas varied by histology; osteosarcomas (Osteosarc) being associated with a high activity of APOBEC and dMMR signatures, leiomyosarcomas are enriched in the UV signature SBS7a and liposarcomas in the chromothripsis associated amplification signatures CN8, SV4 and SV6^31^. As expected colon cancers show evidence of dMMR (SB15, SBS26, SBS44)^32^ whereas rectal cancers tend to feature SBS88, indicative of colibactin exposure as a consequence of pks^+^ E.coli infection^33^. Clock-like signature activity (e.g. SBS1 and SBS40) is higher in IDH wild-type glioblastomas (GBM-IDHwt) compared to mutated (GBM-IDHmut), which reflects older age at diagnosis. Similar relationships are apparent in other cancer types. In lung cancer, squamous cell carcinomas (SCC) are enriched for APOBEC signatures (SB2, SBS13) and small cell tumours for the smoking signature SBS92^34, 35^. Among haematological malignancies, multiple myeloma (MM) and ALL have the highest mutation rates with APOBEC signatures particularly active in MM likely as a consequence of altered APOBEC3G activity^36^.

**Figure 4.**
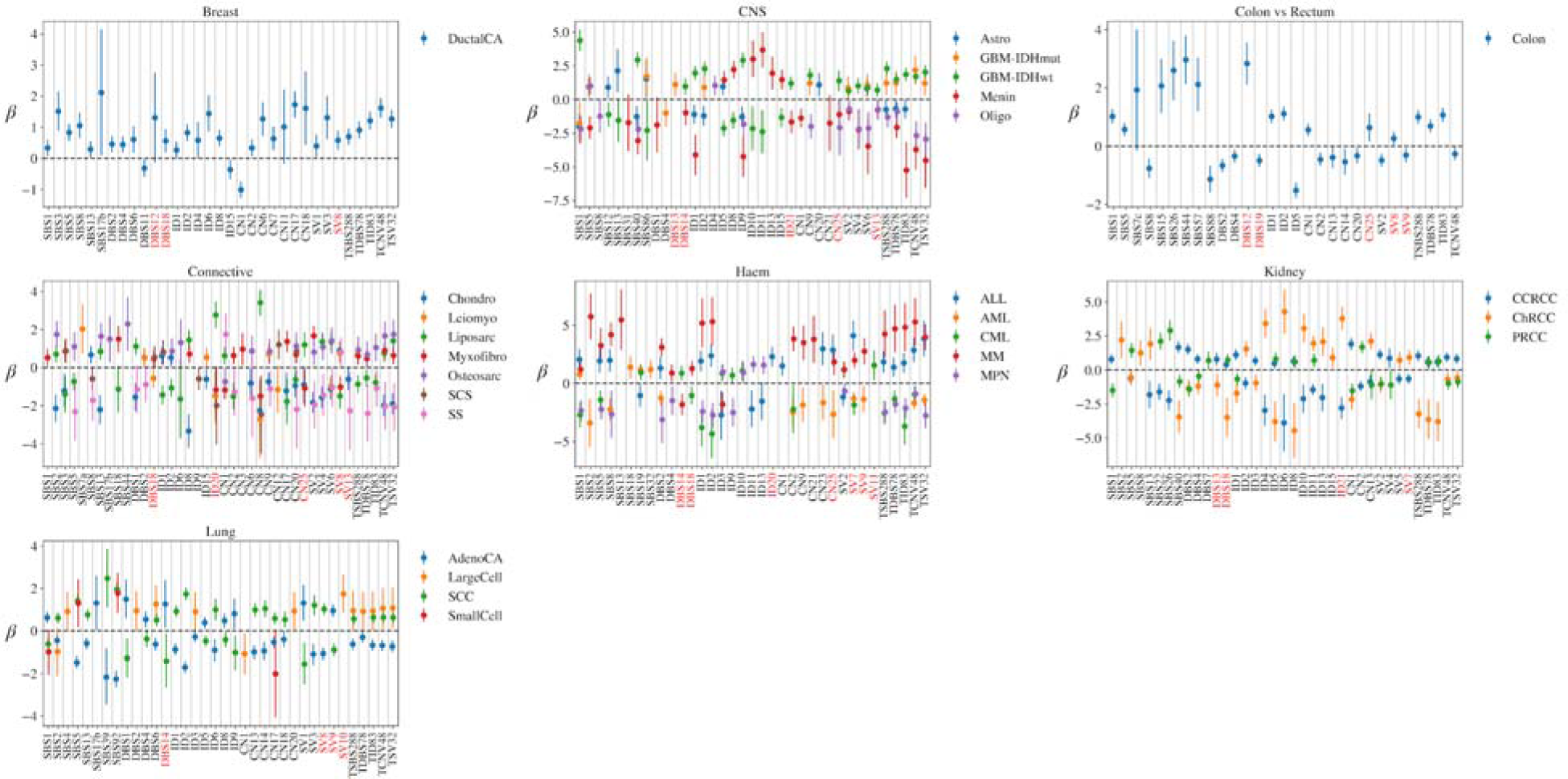
Association between signature activity and tumour histology within each cancer type. The logistic regression coefficient of the signatures being active (or having activity greater than the median) for a tumour having the given histology compared with the remainder of the cohort. Error bars show the 95% confidence interval for the regression coefficient and cohort descriptions with abbreviations are listed in **Supp. Table 3**.

We identified a number of significant signature-DNA repair gene and signature-treatment associations (FDR<0.01, **Methods Page 11**, **Supp. Materials Page 3**). These include known associations such as dMMR signatures (e.g. SBS44) with *MSH6* inactivation in CRC (*P*-value: *P*=3.5e-62, effect size: ES=1.12), *POLE* signatures (e.g. SBS10a) associated with *POLE*-inactivation in uterine cancer (*P*=2.1e-15, ES=2.47), and DBS5 with oxaliplatin exposure in CRC (*P*=5.3e-47, ES=2.9). We also identified relationships between signatures, genes and treatments that have not been previously reported (**Fig. 5**). Signatures SBS93, DBS4 and ID14, which have correlated activities across samples, are associated with *POLG* inactivation in colorectal cancers (*P*=1.3e-9, 4.0e-7, 7.9e-4, ES=0.4, 0.2, 0.3). SBS17a, SBS17b and DBS4, three other signatures that also have correlated activity, are significantly associated with *POLG* inactivation under logistic regression (*P*=5.2e-4, 2.7e-5, 2.2e-6, beta=0.4, 0.4, 0.4). The novel signature CN25 is associated with *MSH6* inactivation in CRC suggesting that this copy number signature is related to dMMR (*P*=5.1e-6, ES=0.7). ID8, a signature of NHEJ, is significantly associated with radiotherapy exposure in both GBM-IDHwt and head and neck squamous cell carcinomas (*P*=1.8e-22, 3.9e-13, ES=3.8, 1.8) even when only primary tumours are considered (removing primary recurrences, *P*=3.9e-10, 3.9e-15, ES=3.5, 1.9), which supports hypotheses that radiation can generate double strand breaks repaired by NHEJ. ID5 is associated with radiotherapy in GBM-IDHwt, CRC, breast lobular cancer and oligodendroglioma (*P*=8.9e-13, 1.5e-8, 7.9e-5, 3.5e-4, ES=2.7, 1.0, 1.3, 2.0) implying radiation exposure induces single base pair A/T deletions.

**Figure 5.**
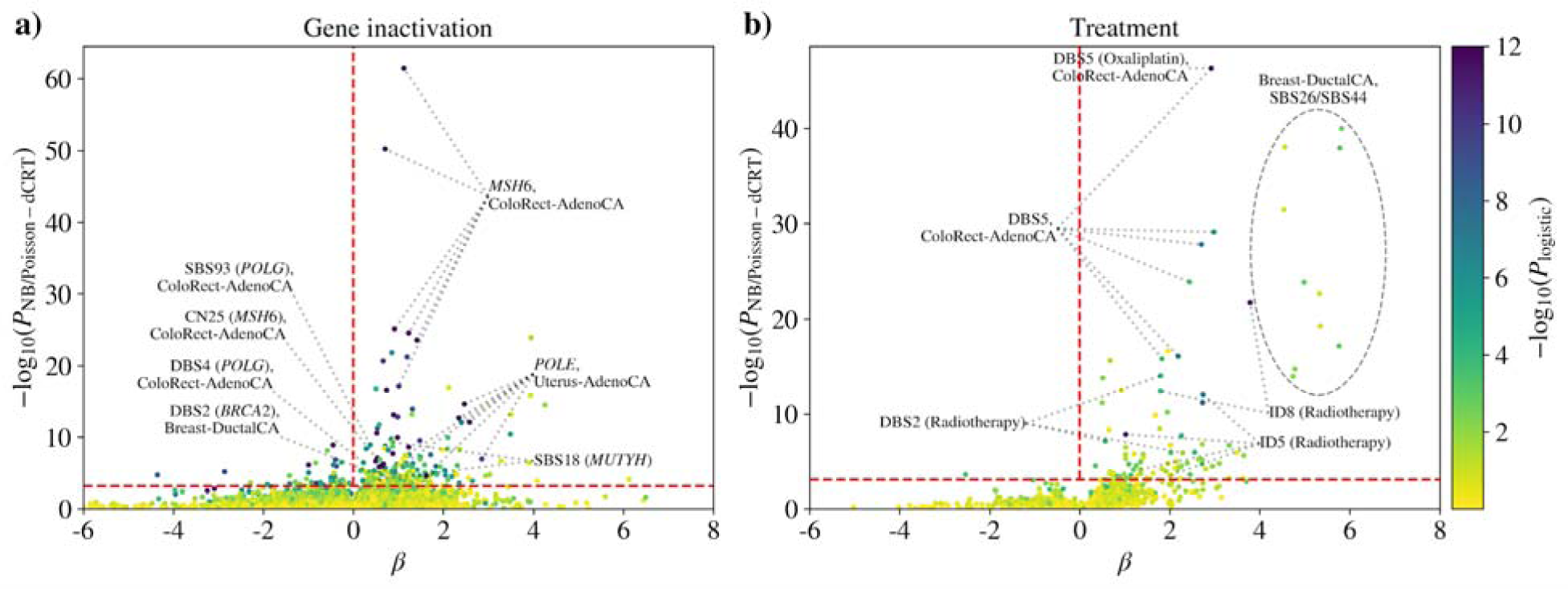
Relationship between DNA repair gene inactivation or treatment exposure and mutational signatures. Associations are computed with the gene knockout parameter for each combination of signature, gene and tumour type, using both a negative binomial and logistic regression linear models. The negative binomial *P*-values are computed using conditional resampling and shown on the y-axis. The logistic regression *P*-values are computed using a Wilks likelihood ratio test and are shown as point colours. **(a)** The majority of significant signature-gene inactivation results have positive association coefficients as expected (**Supp.** Fig. 5), most notably *MSH6* inactivation in colorectal cancer and *POLE* gene inactivation in uterine cancers. The horizontal dashed line corresponds to an FDR of 0.01. **(b)** Treatment exposures are also associated with an increase in signature activities.

Many of the other associations we identified may be a consequence of gene inactivations informing treatment decisions. For example, 67% (20/30) of breast ductal cancer patients who received 5-Fluorouracil (5-FU) treatment have at least one *MLH1* allele inactivated compared with 28% of the whole cohort (Fisher exact *P*-value=1.2e-5). Inactivation of *MLH1* increases dMMR signature activities resulting in associations between SBS26 or SBS44 and 5-FU or related treatments (**Fig. 5b**). All associations of signatures with gene inactivations and treatment exposures are given in **Supp. Tables 6 and 7** respectively.

**Fig. 6** shows the relationship between signature activities and tumour stage, grade and hormone status of breast cancers. As previously documented, HRD signatures are associated with tumour grade and are inflated in ER and PR-negative breast ductal cancers^37^. However, HER2 status does not show a strong relationship with HRD and HER2-positive breast ductal cancers tend to have higher rates of APOBEC-associated signatures^38, 39^ (**Fig. 6b,d**). HER2 promotes cell growth and is associated with more aggressive tumours^40^ and APOBEC activity is significantly associated with overall survival (**Fig. 7b**). dMMR signature activity is inflated in CRC with higher grades but lower stage (**Fig. 6c**) and a similar relationship is seen with *POLE* signatures in colorectal and uterine cancers.

**Figure 6.**
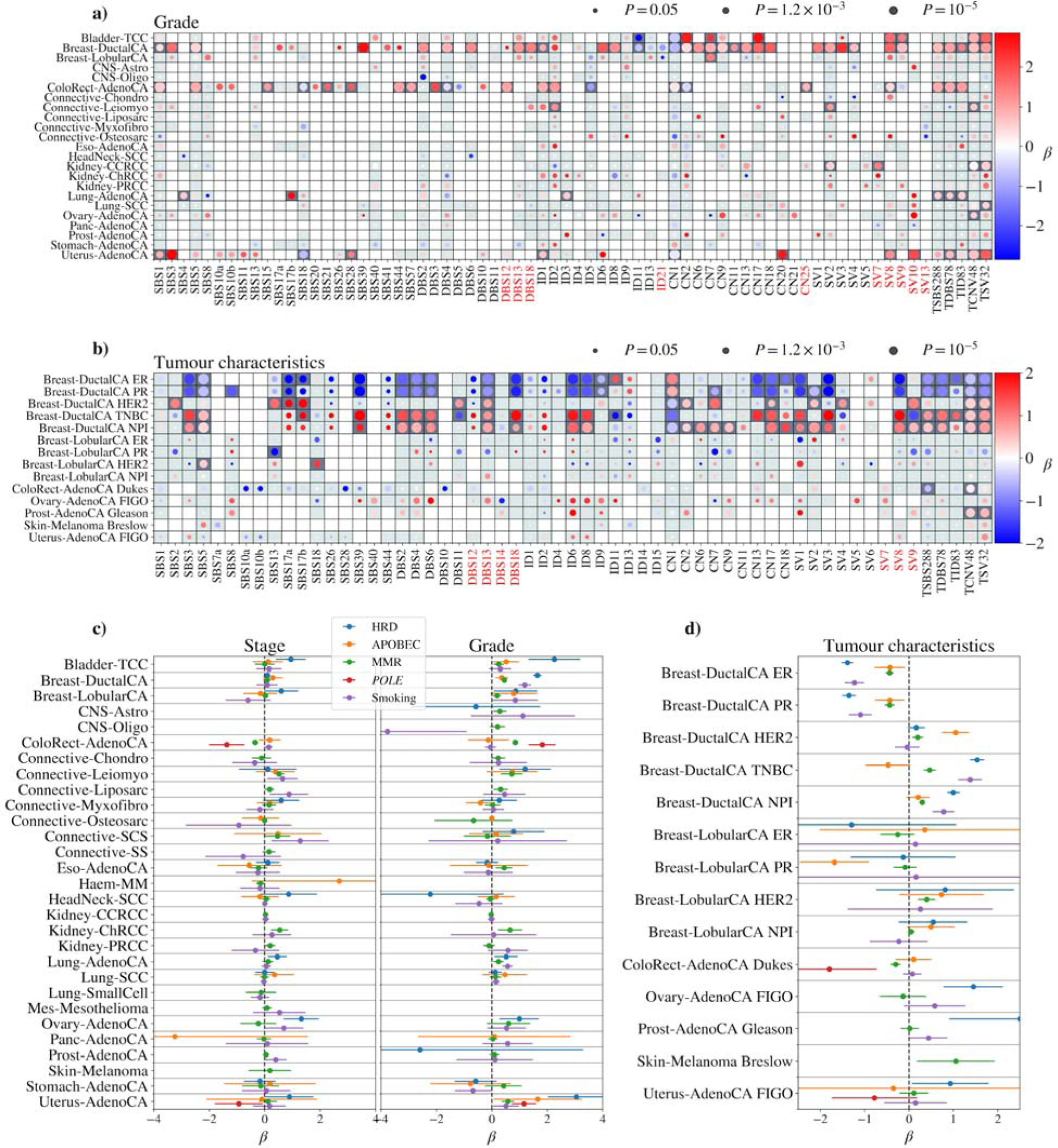
Relationship between mutational signatures and tumour histology. Logistic regression is performed on signature activities (either non-zero or above median) with clinical properties of tumours as covariates in tumour type groups. **(a)** Associations with tumour grade from PHE/NCRAS where the size of the bubble reflects the Wilks *P*-value of association and colours show the association coefficient. **(b)** As in **(a)** but for tumour-type specific features such as hormone status and relevant tumour grades. **(c)** Signatures with known processes are grouped together and inverse-variance weighted posteriors are shown for the association of tumour TNM stage and grade in each cohort. **d)** As in **(c)** but for the tumour type-specific features.

**Figure 7.**
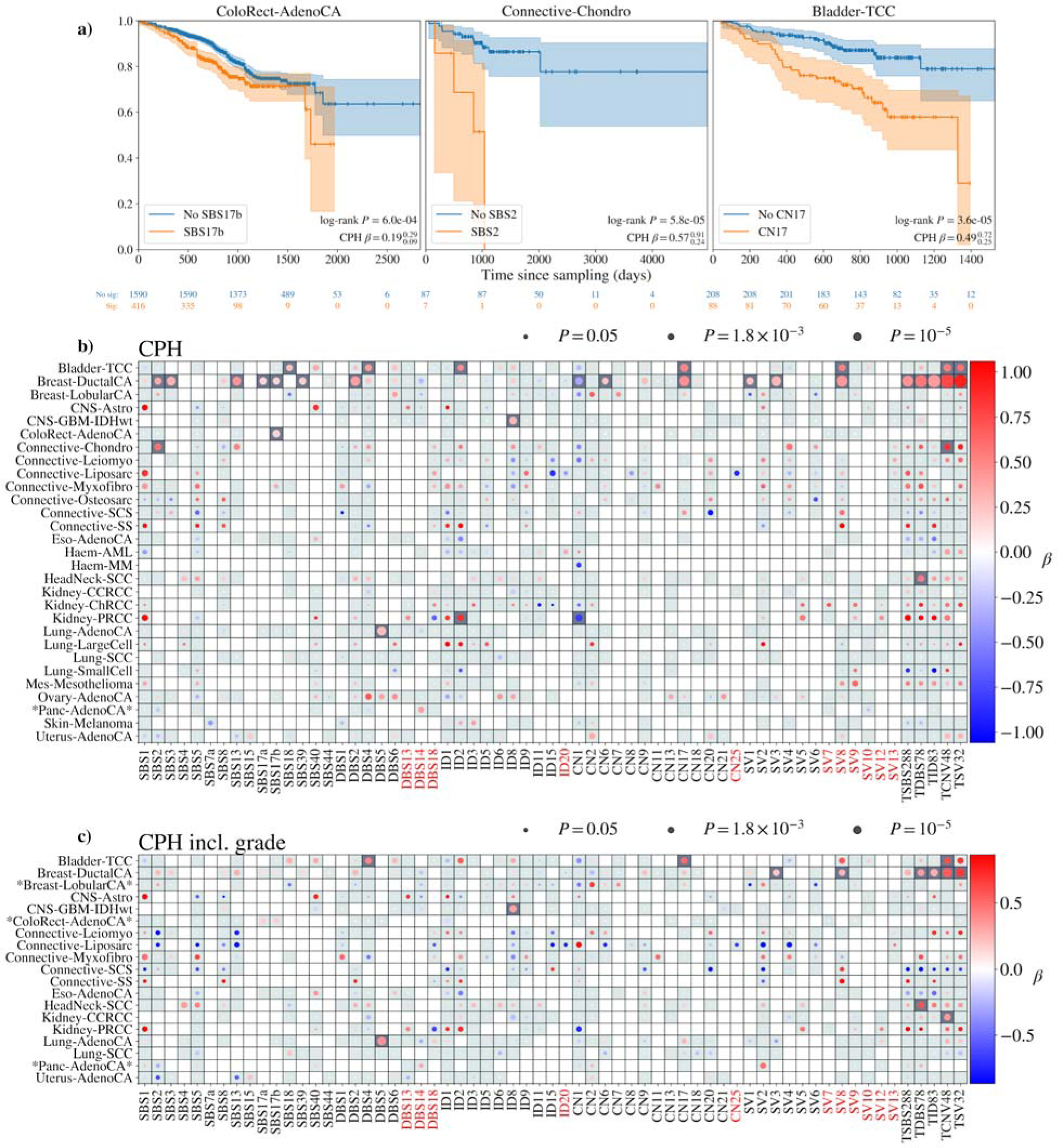
Relationship between mutational signatures and patient overall survival. **(a)** Survival KM curves for colorectal, chondrosarcoma and transitional cell carcinomas for samples with SBS17b, SBS2 and CN17 active respectively. In all cases the presence of the signature reduces the expected survival time of the patients. **(b)** Cox proportional hazard tests are performed for each signature in each tumour type. Positive association indicates a reduced survival for participants with that signature active in the tumour. Patients with breast ductal cancer have particularly worse survival where HRD signatures are active. **(c)** As per figure **(b)** but with grade of cancer included as a covariate.

### Prognostic and therapeutic insights from mutational signatures

Specific signatures are associated with survivorship in a number of tumour types (**Fig. 7**). Notably, HRD and APOBEC-related signatures in ductal breast cancers are associated with a poorer patient outcome even after adjusting for tumour grade. Signature SBS17b is associated with reduced CRC survival (*P*=6.0e-4, Cox PH beta=0.19, 95% CI [0.09, 0.29]). APOBEC activity indicated by SBS2 is associated with reduced survival of chondrosarcoma (*P*=5.8e-5, beta=0.57, [0.24, 0.91]). CN17, active in 88 out of 296 bladder transitional cell carcinoma (TCC), is significantly associated with reduced survival (*P*=3.6e-5, beta=0.49, [0.19, 0.80]). While DBS5 is associated with reduced survival in lung adenocarcinoma and ovarian cancer this is likely to be the consequence of confounding by indication, since DBS5 can be a consequence of platinum therapy. In GBM-IDHwt, a higher ID8 burden, reflective of NHEJ inactivation, is associated with reduced overall survival, however, this is caused by recurrent tumours with worse survival which also have significantly increased ID8 burden. The signal is significantly reduced when only considering primary tumours.

Mutational signatures also indicate the DNA repair capacity of cancer cells, and as such are increasingly being shown to predict response to DNA-damaging or other therapies^41^. Notably HRD signatures provide an indication for PARP inhibition therapy^42^. There is also increasing evidence that HRD status may indicate sensitivity to platinum chemotherapy^43^. Using non-zero activity in at least two of SBS3, ID6 and CN17 in a sample as an indicator of HRD, we find that 269 breast cancers, 132 ovarian, 41 lung, 31 uterus, 28 connective tissue and 23 cancers from other tissue types show evidence of HRD. The aetiological basis of HRD is identifiable in 16% cases on the basis of biallelic inactivation of *BRCA1, BRCA2, PALB2, BRIP1* or *RAD51B* through germline and somatic mutations. Many of the other cases may be caused by promoter methylation; however, this data is not available for 100kGP samples. Our analysis suggests that many more patients may benefit from HRD-targeting therapies than are currently eligible based on driver gene identification.

While high levels of dMMR associated signatures are primarily a feature of uterine (35%) and colorectal (18%) cancer, it is also seen in subsets of other tumours including those of the lung, ovarian, prostate, kidney and breast. However, only 11% of dMMR tumours show evidence of *MSH6*, *MSH2*, or *MLH1* inactivation. The identification of dMMR signatures has therapeutic relevance: for solid tumours is means eligibility for checkpoint inhibition^44^; sensitivity to 5-FU^45^ and Werner syndrome helicase (WRN) inhibition^46, 47^ and indicates gliomas which may respond adversely to temozolomide and thiopurine therapy due to mutagenesis of driver genes such as *TP53*^41^.

Finally, APOBEC signatures SBS2 and SBS13 are both present in 13% breast, 10% ovarian, 9% connective and 7% of uterus and stomach cancers. APOBEC signatures are associated with sensitivity to ATR inhibition in multiple cancer cell lines including breast and ovarian^48^.

## DISCUSSION

We extracted mutational and chromosomal alteration signatures from 10,983 tumour samples across 16 tumour types as part of the 100,000 Genomes Project. This is the first study to extract and analyse the full complement of SBS, DBS, ID, CN and SV signatures from a single cohort and the largest study of ID, CN and SV signatures to date. 29 of our extracted signatures are not currently listed on COSMIC, 7 of which (DBS19, ID19-22, CN25 and SV12) are linearly independent and have not been seen in any previous studies.

Degasperi *et al*.^16^ have also extracted SBS and DBS signatures in 100kGP data. There are salient differences between our analysis and Degasperi *et al*. which impact on recovery of signatures. In particular, we excluded samples with incorrectly recorded histologies and PCR-amplified samples, and only claimed novel signatures where they were linearly independent of pre-existing COSMIC signatures. This is discussed further in the **Page 4 of Supplementary Materials** where we demonstrate that almost all signatures reported as novel in Degasperi *et al.* can be constructed from linear combinations of signatures in COSMIC and this work.

Using the curated histology data we compared signature activities between tumour groups and found signatures corresponding to processes that are more prevalent in some tumour types than others. For example, SBS22 has previously been linked to exposure to the nephrotoxic aristolochic acid. Herein we demonstrated a relationship between SBS22 and renal cancer, with specificity for papillary subtype. We have found associations between signatures with hitherto unknown aetiologies and DNA repair gene inactivation or treatment exposures. Notably, the novel signature CN25 is associated with *MSH6* inactivation and as such is a signature of dMMR.

For the first time in a study of this scale, we have analysed the implications of signature activities for patient outcome. Our analysis shows that signatures can additionally inform patient prognosis above and beyond conventional clinical staging. Signatures of HRD, dMMR and APOBEC activity are also indicators for patient response to multiple therapies including immune checkpoint inhibitors. However, if signature analysis is to become part of routine patient care as envisaged by the 100kGP it is essential that analyses should be cognisant of the statistical issues surrounding assignment of signatures and potential errors both from sequencing artefacts and downstream analyses. As well as providing additional insights into tumour biology our analysis shows the relevance of signature analysis for delivering the vision of precision oncology.

## Data Availability

All data produced are contained in the supplementary tables. Any data which could not be provided in the tables is available through the Genomics England research environment.

## Code availability

**Supp. Table 11** lists software used in this work and SigProfilerExtractor parameters used for signature extraction.

## Data availability

All WGS data and processed files from the 100,000 Genomes Project can be accessed by joining the Pan Cancer Genomics England Clinical Interpretation Partnership (GeCIP) Domain once an individual’s data access has been approved (https://www.genomicsengland.co.uk/portfolio/pan-cancer-across-cancers-gecip-domain/). The link to becoming a member of GECIP and having access can be found here https://www.genomicsengland.co.uk/research/academic/join-gecip. The process involves an online application, verification by the applicant’s institution, completion of a short information governance training course, and verification of approval by Genomics England. Please see https://www.genomicsengland.co.uk/research/academic for more information. The Genomics England data access agreement can be obtained from https://figshare.com/articles/GenomicEnglandProtocol_pdf/4530893/5. All analysis of Genomics England data must take place within the Genomics England Research Environment (https://www.genomicsengland.co.uk/understanding-genomics/data). The 100,000 Genomes Project publication policies can be obtained from https://www.genomicsengland.co.uk/about-gecip/publications.

## Supplementary Materials

**Supp. Table 1**: Selection of tumour samples from the 100,000 Genomes Project pancancer domain

**Supp. Table 2**: Tumour groups included in study

**Supp. Table 3**: Signatures extracted, possible decompositions and previously extracted signatures with high similarity

**Supp. Table 4a-e**: Mutational signature activities in tumour samples for SBS, DBS, ID, CN and SV mutation classes

**Supp. Table 5a-e:** Mutational signatures for SBS, DBS, ID, CN and SV mutation classes

**Supp. Table 6**: Association statistics between signature activities and tumour suppressor gene inactivation

**Supp. Table 7**: Association statistics between signature activities and treatment received by participants

**Supp. Table 8**: Association statistics between signature activities and clinico-pathological features

**Supp. Table 9**: Association between signature activities and tumour histology

**Supp. Table 10**: Survival analysis

**Supp. Table 11:** Software and SigProfilerExtractor parameters used for signature extraction of each mutation type

## GENOMICS ENGLAND RESEARCH CONSORTIUM

Ambrose, J. C.^1^; Arumugam, P.^1^; Bevers, R.^1^; Bleda, M.^1^; Boardman-Pretty, F.^1, 2^; Boustred, C. R.^1^; Brittain, H.^1^; Brown, M.A.; Caulfield, M. J.^1, 2^; Chan, G. C.^1^; Giess A.^1^; Griffin, J. N.; Hamblin, A.^1^; Henderson, S.^1, 2^; Hubbard, T. J. P.^1^; Jackson, R.^1^; Jones, L. J.^1, 2^; Kasperaviciute, D.^1, 2^; Kayikci, M.^1^; Kousathanas, A.^1^; Lahnstein, L.^1^; Lakey, A.; Leigh, S. E. A.^1^; Leong, I. U. S.^1^; Lopez, F. J.^1^; Maleady-Crowe, F.^1^; McEntagart, M.^1^; Minneci F.^1^; Mitchell, J.^1^; Moutsianas, L.^1, 2^; Mueller, M.^1, 2^; Murugaesu, N.^1^; Need, A. C.^1, 2^; O‘Donovan P.^1^; Odhams, C. A.^1^; Patch, C.^1, 2^; Perez-Gil, D.^1^; Pereira, M. B.^1^; Pullinger, J.^1^; Rahim, T.^1^; Rendon, A.^1^; Rogers, T.^1^; Savage, K.^1^; Sawant, K.^1^; Scott, R. H.^1^; Siddiq, A.^1^; Sieghart, A.^1^; Smith, S. C.^1^; Sosinsky, A.^1,2^; Stuckey, A.^1^; Tanguy M.^1^; Taylor Tavares, A. L.^1^; Thomas, E. R. A.^1,2^; Thompson, S. R.^1^; Tucci, A.^1,2^; Welland, M. J.^1^; Williams, E.^1^; Witkowska, K.^1,2^; Wood, S. M.^1,2^; Zarowiecki, M.^1^.

1. Genomics England, London, UK.

2. William Harvey Research Institute, Queen Mary University of London, London, UK.

## ACKNOWLEDGEMENTS

This research was made possible through access to the data and findings generated by the 100,000 Genomes Project. The 100,000 Genomes Project is managed by Genomics England Limited (a wholly owned company of the Department of Health). The 100,000 Genomes Project is funded by the National Institute for Health Research and NHS England. The Wellcome Trust, Cancer Research UK and the Medical Research Council have also funded research infrastructure. The 100,000 Genomes Project uses data provided by patients and collected by the National Health Service as part of their care and support. R.S.H. acknowledges support from the Wellcome Trust (214388) and Cancer Research UK (C1298/A8362). A.S. and J.J. are in receipt of a National Institute for Health Research (NIHR) Academic Clinical Lectureships, funding from the Royal Marsden Biomedical Research Centre, a starter grant for clinical lecturers from the Academy of Medical Sciences. D.N.C. and A.F. are funded by the Oxford NIHR Comprehensive Biomedical Research Centre (BRC). D.C.W. is funded by the NIHR Manchester Biomedical Research Centre. A.E. is grateful to Eugene Katsevic and Tim Barry for their help understanding the conditional resampling method which was generalised to signature analysis in this work.

## AUTHOR CONTRIBUTIONS

A.J.C and D.Chubb processed clinical data. A.J.C, D.Chubb, A.G and A.E performed quality control on SBS, DBS and ID somatic mutations. A.J.C and A.H called copy number alterations. A.J.C called structural variants. D.Chubb, A.J.C, A.J.G, A.H and A.T optimised SigProfilerExtractor. A.J.G and M.B extracted SBS and DBS signatures, A.J.G, M.B and A.E extracted ID signatures, A.H. extracted CN signatures and A.T extracted SV signatures. A.E combined signatures pancancer. A.E analysed signatures against clinical variables, gene knockouts, treatments and survival. A.E and D.Chubb analysed this work against Degasperi *et al* 2022. D.C.W, A.J.G. and R.S.H supervised the study. A.J.C, A.E, A.H, A.J.G, A.T, A.S, D.C.W and R.S.H wrote the manuscript. All authors read and approved the final version of this manuscript.

