## Supplementary material for "Comprehensive repertoire of the chromosomal alteration and mutational signatures across 16 cancer types from 10,983 cancer patients": Methods

Online methods

### 100,000 Genomes Project whole genome sequencing data

14,129 paired tumour and germline samples were obtained from V11 of the GEL 100kGP. Samples were prepared using Illumina TruSeq DNA PCR-free library preparation kit and sequencing performed on a HiSeq X producing 150 base pair (bp) paired-end reads to 33x depth for germline and 100x for tumour samples. The 100kCP program excludes outliers with poor sequencing quality (based on the percentage of mapped reads, percentage of chimeric DNA fragments, average insert size, AT/CG dropout and unevenness of local coverage). Alignment was performed to the *Homo sapiens* GRCh38decoy assembly using Isaac v03.16.02.19 (Raczy *et al.* 2013).

### Collection and processing of clinical data

Clinical and demographic data were obtained from NHS Digital (NHSD), Public Health England’s National Cancer Registration and Analysis Service (PHE-NCRAS) and the Genomic Medicine Centers (GMCs) through the Genomics England Research Environment. Additional data were also obtained from associated histology reports where available. Sequenced tumour samples were matched to their respective PHE-NCRAS records using the tumour sampling date and PHE-NCRAS treatment dates, allowing a maximum discrepancy of 28 days. Obtained data comprised sex, year of birth, date of cancer diagnosis, date of last reported clinical follow-up, survival outcome and date of death if relevant, tumour histology, anatomical site of primary tumour, anatomical site sampled, and whether the sample was taken from a primary tumour, a metastasis or a recurrence of a primary tumour. For some variables data was obtained from multiple sources (GMC, NHSD, PHE-NCRAS). Any potential conflicts between these data sources were reviewed manually. Where possible, sequenced tumours were assigned to one of 41 tumour groups depending on the tumour histology and originating tissue (Supplementary Table 3). The 41 tumour groups were chosen based on availability of sequenced samples and prior knowledge of the genomic similarity of disease subtypes.

Whether participants had received systemic treatment or disease-associated radiotherapy prior to sampling was determined using data from NHSD and PHE-NCRAS. Admitted patient care and outpatient records related to systemic treatment were obtained from NHSD tables using Office of Population Censuses and Surveys (OPCS)-4 codes. Records related to systemic treatment were also obtained from the PHE-NCRAS AV Treatment and Systemic Anti-Cancer Therapy (SACT) tables. Records related to radiotherapy were obtained from the PHE-NCRAS AV Treatment and National Radiotherapy Dataset (RTDS) tables.

### Calling small germline and somatic variants

Calling of germline SNPs and indels was performed using Starling (version 2.4.7) (Raczy, *et al*., 2013). Calling of somatic SNVs and indels was performed using Strelka (version 2.4.7) (Kim, *et al*., 2018). In addition to the default Strelka filters, somatic variants were removed if they met any of the following criteria:

- Had a population germline allele frequency ≥1% in the 100KGP or gnomAD cohorts (Karczewski, *et al*., 2020).
- Had a somatic frequency ≥5% in the 100kGP tumour samples.
- Overlapped a simple repeat as defined by Tandem Repeats Finder (Benson, 1999).
- Was an SNV likely resulting from systematic mapping or calling artefacts. Likely artefacts were identified by computing the ratio of tumour allele depths at each somatic SNV site and comparing against the ratio of allele depths at the same site in a panel of normal samples, which comprised 7,000 non-tumour genomes from the 100kGP cohort. Allele depths at each site were counted in the panel of normal, including only individuals not carrying the relevant alternate allele. To replicate Strelka filters, duplicated reads were removed prior to counting and mapping quality ≥5 and base quality ≥5 thresholds were applied. SNVs with a Fisher’s exact test Phred quality score ≤50 were excluded.
- Was an indel in a region of high levels of sequencing noise, where ≥10% of the base calls in a window extending 50 base pairs to either side of the indel call have been filtered out by Strelka.
- Was an InDel within 10bp of an InDel called in GEL or in GNOMAD in >1% of germline samples.

### Calling somatic copy number alterations

Somatic copy number alterations (CNAs) were called using a five-stage procedure (Nik-Zainal, *et al*., 2012):

*Stage I: Initial copy number alteration profiling*

Clonal and sub-clonal CNAs were profiled using Battenberg (Nik-Zainal *et al.* 2012). Briefly, alleleCount-FixVAF was used to count reads supporting single-nucleotide polymorphism (SNP) reference and alternate alleles (Cornish *et al.* 2020). Heterozygous SNPs were then phased with SHAPEIT2 (version 2.r904) (Delaneau *et al.* 2011). Piecewise constant fitting was used to segment phased SNPs (Nilsen *et al.* 2012) and CNAs with evidence of subclonality were identified using t-tests. Sample tumour purity and ploidy were estimated using the approach described by Van Loo *et al.* (2010). Sequencing data were aligned to hg38 and it was therefore necessary to convert SNP positions to hg37 before phasing and convert output segments back to hg38.

*Stage II: Using variant allele frequency distributions to evaluate profile concordance*

Factors influencing expected variant allele frequencies (VAFs) include (a) the fraction of tumour cells containing the variant, (b) CNAs at the variant site, (c) the number of chromosome copies carrying the variant (multiplicity) and (d) tumour sample purity (Dentro *et al.* 2017). Given the tumour copy number profile and sample tumour purity we can expect to observe enrichment of variants with VAFs approximating specific values, representing clonal variants present in all tumour cells (Van Loo *et al.* 2010). Failure to observe this enrichment indicates that either the copy number profile or tumour sample purity is incorrect. We therefore used SNV VAF distributions to assess the CNA profiles and sample tumour purities computed by Battenberg (Househam *et al.* 2022).

Autosomal genome segments with copy number states of 1:1, 1:0, 2:2, 2:1, 2:0 with no evidence of subclonal CNAs were considered when assessing SNV VAF distributions. The five copy number states were considered separately as expected clonal SNV VAFs and possible variant multiplicities differ between states (Van Loo *et al.* 2010). A copy number state was not considered if it corresponded to genome regions containing <5% of all SNVs. Expected VAF distribution peak locations were computed as:

$$\frac{\rho_{Battenberg}m}{2\left( 1-\rho_{Battenberg} \right)+\rho_{Battenberg}\psi_{v}}$$

Where ρ_Battenberg_ is the sample tumour purity output by Battenberg, ψ*_v_* is the tumour ploidy at the variant site, and *m* is the variant multiplicity (which can equal 1 or 2 in 2:2, 2:1 and 2:0 states and only 1 in 1:1 and 1:0 states). VAF distribution peaks were identified using kernel density estimation implemented in the peakPick R package (version 0.11) (Weber *et al.* 2014). Peaks with densities <0.3 were excluded. For each copy number state, the expected peak location corresponding to the greatest variant multiplicity was matched to the observed VAF distribution peak with the greatest VAF. Remaining expected peak locations were then matched to the observed peaks with most similar VAFs. Tumour heterogeneity can inhibit VAF peak detection and therefore for samples where ≥1 expected peaks were considered, the expected peak furthest from the respective matched observed peak (in terms of VAF) was discarded. Sample tumour purity (ρ_i_) was then estimated for each remaining expected peak using its matched observed peak VAF:

$$\rho_{i}=\frac{2a}{m+\omega(2-\psi_{s})}$$

where ω is the VAF of the matched observed peak and ψ*_s_* is the state ploidy. A single new purity estimate (ρ_new_) was then computed as a weighted average of the peak-wise purity estimates:

$$\rho_{new}=\sum_{i} \frac{{n_{i}\rho}_{i}}{Nq_{i}}$$

where q_i_ is the number of considered variant multiplicities for the copy number state, n_i_ is the number of SNVs in genome regions with the copy number state, and N is the number of SNVs in genome regions of all considered copy number states. Finally, the difference between the Battenberg purity estimate and the peak-wise purity estimates was used to assess CNA profile quality:

$\eta=\sum_{i} \frac{n_{i}|\rho_{i}-\rho_{Battenberg}|}{Nq_{i}}$.

*Stage III: Profile quality assessment*

The following criteria were used to assess CNA profile quality:

- SNV VAF distribution peaks found at expected locations (defined as η<5%).
- A clonal variant cluster was identified by DPClust (Nik-Zainal *et al.* 2012). This was defined as a variant cluster with a cancer cell fraction (CCF) between 0.9 and 1.1 containing ≥5% of all SNVs.
- No “super-clonal” variant clusters were identified by DPClust. These were defined as variant clusters with CCFs>1.1 containing ≥5% of all SNVs.
- If most of the genome is categorised as 2:2 (tetraploid) then a SNV VAF distribution peak in 2:2 regions corresponding to a variant multiplicity of 1 was observed.
- No homozygous deletions >10Mb are called.

CNA profiles not satisfying at least one criterion were deemed to fail and were re-profiled (*i.e.* proceeded to Stage IV). CNA profiles satisfying all criteria were deemed to pass and were used in subsequent analyses.

*Stage IV: Copy number alteration re-profiling*

Samples failing CNA profile quality assessment were re-profiled a maximum of three times using alternative tumour sample purity and ploidy estimates. CNA profiles still failing quality assessment after three attempts were not considered in later analyses. New purities (ρ_new_) were iteratively re-estimated as per Stage II, whilst new ploidies (ψ_new_) were estimated as per Van Loo *et al.* (2010):

$$\psi_{new}=\frac{\rho_{Battenberg} \left( \psi_{Battenberg}-2 \right)+2\rho_{new}}{\rho_{new}}$$

*Stage V: Manual review*

Three tumour groups (Haem-MPN, Ovary-AdenoCA and Testis-GCT) had higher than expected sample proportions failing CNA profile quality assessment. High failure rates in Haem-MPN and Testis-GCT were due to low SNV, which complicated automatic identification of SNV VAF distribution peaks. Conversely, the high failure rate in Ovary-AdenoCA was primarily due to the presence of large homozygous deletions. For these three tumour groups we therefore manually reviewed CNA profiles failing quality assessment and passed them when appropriate. The biological plausibility of large homozygous deletions in Ovary-AdenoCA tumours was assessed by considering the involvement of genes classified as essential in ovarian cancer cell lines in DepMap (Tsherniak *et al.* 2017). If a homozygous deletion contained no essential genes, then it was considered biologically plausible, and the CNA profile was considered passable.

### Calling and classifying somatic structural variants

Somatic structural variants (SV) were called using Delly (Rausch *et al.* 2012), Lumpy (Layer *et al.* 2014) and Manta (Chen *et al.* 2016) with a graph-based consensus approach, with support from CNA profiles. SVs were first called using the three SV callers, with default parameters. Delly was run with post-filtering of somatic SVs using all normal samples. SVs from the individual callers were removed if: (a) any reads supporting the variant were identified in the matched normal sample, (b) <2% tumour reads supported the variant, (c) either variant breakpoint was located on a non-standard reference contig (not chromosomes 1-22, X or Y), or (d) either variant breakpoint was located in a centromeric or telomeric region. Remaining SVs were merged with a modified version of PCAWG Merge SV, allowing 400bp slop at the breakpoint positions (Li *et al.* 2020). SVs were included in the final SV data set if they were supported by at least two of the three SVs callers, or by only one SV caller but with a breakpoint <3kb from a CNA segment boundary.

xTea was used to call somatically acquired long interspersed nuclear element (LINE-1) retrotransposition events (Chu *et al.* 2021). Alu elements, SINE-VNTR-Alu elements and processed pseudogenes comprise ≤3% of cancer retrotransposition events and were therefore not analysed (Rodriguez-Martin *et al.* 2020). Retrotransposition events were not considered in subsequent SV analyses as they are mechanistically distinct from other SV-generating events (Rodriguez-Martin *et al.* 2020). SVs were categorised as likely retrotransposition events and excluded if: (a) a transduced region was identified in the same tumour sample within 10kb of either rearrangement breakpoint, or (b) a transduced region was identified within 10kb of either rearrangement breakpoint in ≥1% tumour samples. A threshold of 10kb was used as the majority of somatically acquired transductions comprise regions <10kb from a LINE-1 element (Tubio *et al.* 2014).

Using ClusterSV^20^, rearrangements were grouped into footprints and clusters based on their proximity within the genome, rearrangement size, and overall rearrangement number in the genome. Rearrangement footprints represent sets of rearrangement breakpoints that are positionally associated. Rearrangement clusters represent sets of rearrangements that are mechanistically associated and were classified as being a simple (deletions, tandem duplications, balanced inversions, balanced and unbalanced translocations, and simple unclassified events) or complex (chromoplexy, chromothripsis, and complex unclassified events) event. Rearrangement clusters comprising ≤2 or ≥3 individual rearrangements were defined as simple and complex events respectively.

Rearrangement clusters were defined as a chromothripsis event if they met the following criteria:

- At least six interleaved intra-chromosomal rearrangements.
- A contiguous series of four genome segments oscillating between two copy number states, or five genome segments oscillating between three copy number states.
- No evidence that the distribution of intra-chromosomal fragment join orientations diverge from a distribution with equal probabilities for the 4 orientation categories (duplication-like, deletion-like, head-to-head inversion, and tail-to-tail inversion) at a false discovery rate of 0.2.

Rearrangement clusters were defined as a chromoplexy event if they met the following criteria:

- Comprises between 3 and 30 rearrangements.
- Contains a chain of rearrangements spanning at least 3 chromosomes. Chains were defined using a graph-based approach, in which nodes represent breakpoints and are connected by an edge if they fall within 1Mb of each other and are not involved in the same rearrangement.
- At least 50% of rearrangement footprints represent balanced translocations, either with a deletion bridge between the break ends or no observed copy number change.

### Selection of study samples

Sequenced tumour samples were excluded if clinical data were missing or if unresolvable conflicts existed between the clinical data sources (GMCs, NHSD, PHE-NCRAS, histology reports) (Supplementary Table 2). In total 2,251/14,129 (15.9%) of tumour samples were excluded based on the following criteria:

- Sex reported by NHSD, PHE-NCRAS and/or the GMC did not match the sex inferred from the sequencing data.
- Sample could not be assigned to one of the 41 tumour groups, either because of missing or conflicting tumour histology or originating tissue data, or because the disease was not represented by one of the groups.
- Missing or conflicting data meant it was unclear whether a primary tumour, a metastasis or a recurrence of a primary tumour was sampled.
- Missing or conflicting data concerning the day of sampling.
- Participant was less than 18 years old on the day of sampling.

Tumour sample purity and sequencing data quality affect the sensitivity and precision of variant calling (Saunders *et al.* 2012) and we therefore also excluded samples using the following quality control procedures (Supplementary Table 2). In total 267/11878 (2.2%) of tumour samples with required clinical data available were excluded based on sequencing data using the following criteria:

- If cross-contamination of the tumour sample was >1%, as estimated by VerifyBamID (Jun *et al.* 2012).
- If cross-contamination of the matched germline sample was >1%, as estimated by VerifyBamID.
- If the number of SNVs called in a tumour was a low outlier for the assigned tumour group. Outliers were defined as tumours where the Z score of the log number of SNVs was < -3, considering only tumours from the same tumour group.

Duplicate tumour samples were also removed, to ensure that no individual was represented more than one in a tumour group. If multiple sequenced tumour samples from the same tumour group were available for an individual, we preferentially kept primary tumour samples with highest purity, as estimated by Ccube (Yuan *et al.* 2018). Based on these criteria, 10,983 tumour samples were suitable for analysis (Supplementary Table 4). This cohort comprises 10,198 primary tumours, 634 metastases and 151 recurrences of primary tumours from 10,975 individuals. 8 individuals were represented in multiple tumour groups.

### Mutational signature extraction and deconvolution

Integer matrices of mutation counts, with a row for each sample and column for each mutation class, are used as inputs to independent signature extractions. A total of 80 extractions are run across the five mutation types and 16 tumour tissue types.

For the classification of SBS signatures, 96 classes have conventionally been used composed of 6 base substitutions (C>A, C>G, C>T, T>A, T>C, and T>G) and the flanking 5’ and 3’ bases (Alexandrov *et al.* 2013, Alexandrov *et al.* 2020). We extended this scheme to 288 classes by considering the transcriptional context of mutations; whether mutations fell on the transcribed, untranscribed or non-transcribed strand (Islam *et al.* 2022). As per COSMIC, we classified DBS signatures into 78 classes, and small ID signatures into 83 classes according to whether the variant was a deletion or an insertion, variant length, number of reference sequence repeats, and whether the variant is microhomologous. CN signatures were assigned to 48 mutation classes according to length of sequence, CN change and whether there was loss of heterozygosity (LOH) (Steele *et al.* 2022), SVs were assigned to 32 classes based on type and size of the SV and whether it was part of a cluster (Degasperi *et al.* 2020).

Signatures were extracted de-novo using a parallelized version of SigProfilerExtractor (Github version dbb9049, Islam *et al.* 2022). For all signature classes, SigProfilerExtractor was run using random non-negative matrix factorization (NMF) initialization, Gaussian mixture model matrix normalization, 10,000 minimum and 1,000,000 maximum NMF iterations and by minimizing an objective function based on generalized Kullback-Leibler updates. SigProfilerExtractor was applied to each cancer type separately, using between 1 and 25 SBS and ID signatures, between 1-20 DBS and between 1-15 CN and SV signatures. For SBS, DBS, ID and CN signature classes the optimal number of signatures was chosen by considering the average stability across NMF replicates and rank sum tests between acceptable solutions (Alexandrov *et al.* 2020). Deconvolution of SV signatures, especially in breast, ovarian and uterine cancers, recovered multiple single class signatures which is undesirable since single classes are no more informative than the underlying mutation rates. We used the Akaike Information Criterion (AIC) to obtain an optimal solution for SV signatures which reduced the number of single class signatures.

The Akaike Information Criterion (AIC) is an estimate of the evidence of a model given the data. The number of parameters is $K (N + M)$ where $K$ is the number of signatures, N is the number of samples M is the number of mutation types. The mutation rates in channels for each sample are assumed to be Poisson distributed with mean given by the expected mutation rate from the product of signatures ($S$) and activities ($A$)

$AIC = 2K(N+M) - 2\left[ M log\left( S^{T}A \right) - S^{T}A \right]$.

The number of signatures which has the minimum AIC is the model best supported by the data. This downweights solutions with many single mutation class signatures as they don’t provide much more information than the input mutation counts. The AIC for different numbers of signatures is shown in **Supplementary Fig. 1** for SVs in Breast, Uterus and Ovarian cohorts. The best AIC solution can be vastly different to solutions picked by SigProfilerExtractor.

### Signature combiner

Signatures were extracted and decomposed to COSMIC reference signatures (v3.2 for SBS, v3.3 for DBS, ID and CN) in 16 cohorts (**Supplementary Table 2**). However, mutational processes can act in multiple tumour types so we combined the cohort-specific results to generate a single set of pan-cancer signatures.

Firstly all signatures were decomposed into the reference list where possible with cos(sim)>0.8. The cosine similarity between all remaining pairs of novel signatures was calculated. Starting with the highest cosine similarity pair, if cos(sim)>0.8, the signatures were extracted from different cohorts and neither signature is already in a set; a new set was created containing the two signatures. Other novel signatures were added to this set if (i) cos(sim)>0.8 with all members of the set and (ii) they were not extracted in the same cohort as any member of the set. A single signature was then generated to describe this set from the inverse variance weighted mean of signatures in the set

$${S_{j}}^{set} =\frac{1}{\sum_{j=1}^{m} \sum_{i=1}^{n} w_{ij}S_{ij}}\sum_{i=1}^{n} w_{ji}S_{ji}$$

where $w_{ji}$ is the inverse variance of the $j^{th}$ mutation class of signature $i$ from SPE and the signature is normalised. This process was be repeated for all signature pairings with cos(sim)>0.8 producing multiple sets of signatures.

The set with the most members, or, where the number of members was the same, the set with the smallest maximum cosine similarity to any reference signature, was added to the reference list. This produced a new reference list with one additional signature. We then returned to the start and decomposed all signatures to the new reference list. This iterative process continued until all signatures could be decomposed into the reference list with cos(sim)>0.8.

### DNA repair gene inactivation

Associations were tested between signature activities and repair gene inactivating mutations.

A list of DNA repair genes was taken from the overlap between tumour suppressor genes in the COSMIC Cancer Gene Census of known cancer-associated genes (Sondka *et al.* 2018) and the list of genes directly involved in DNA repair mechanisms from <https://www.mdanderson.org/documents/Labs/Wood-Laboratory/human-dna-repair-genes.html#Human%20DNA%20Repair%20Genes> (Wood *et al.* 2001, Knijnenburg *et al.* 2018) (genes labelled as ‘defective in diseases associated with sensitivity to DNA damaging agents’ or ‘other conserved DNA damage response genes’ were not included). This resulted in a list of 41 cancer-associated DNA repair genes.

Germline mutations for each sample in the gene were taken from aggV2 in GEL (https://cnfl.extge.co.uk/pages/viewpage.action?pageId=156601552). Any mutations with CADD>20 (using CADD v1.6, Rentzsch *et al.* 2021) and not classified as “Benign” or “Likely benign” in ClinVar (Landrum *et al.* 2014) were considered as a monoallelic inactivation of one copy of the gene. The distribution of CADD scores for ClinVar variants is shown in **Supplementary Fig. 2** separated by the clinical significance annotation in ClinVar. The threshold of CADD>20 collects the vast majority of pathogenic and only small number of benign variants; however it also selects many missense variants of unknown significance. Therefore the germline mutation rate is higher than is often quoted in the literature from only nonsense and frameshift mutations.

OncoKB (v3.14, Chakravarty *et al.* 2017) was run on somAgg VCFs in GEL to find the number of mutations in a gene for each sample which were classified as “Oncogenic” or “Likely Oncogenic”. Battenberg (Nik-Zainal *et al.* 2012) was used to estimate copy number variants across the genome. If any region overlapping the gene in the sample had LoH in >50% of the tumour sample, this was counted as an inactivation of one allele.

LoH and Germline mutations were treated as binary variables. For many genes, two germline mutations would result in the participant having a congenital condition which would make them unlikely to be a cancer patient in later life in the GEL cohort. Somatic mutations could take any number.

The gene inactivation parameter was the sum of germline, LoH and somatic variants to a maximum of 2 (anything greater than 2 is given the value of 2). Most of the mutation burden was from germline or LoH hits as shown in **Supplementary Fig. 3**, however, somatic mutations are prevalent in cohorts where the gene is a known driver e.g. BRCA2 in ovary or MSH6 in CRC and uterus. In these cases the somatic mutations are likely to be under positive selection for the given tumour type.

### Patient therapy exposure

Chemotherapy and radiotherapy can induce mutations through DNA damage (Kucab *et al.* 2019, Pleasance *et al.* 2020). GEL provides treatment information collected from PHE-NCRAS for many participants which is matched to the tumour samples as described in the clinical data section above. A sample was considered to be exposed to a treatment if the treatment start date was before the sampling date of the tumour. This was only considered as a binary variable. The numbers of patients in each tumour group exposed to each type of therapy is shown in **Supplementary Fig. 8**.

### Associations between signature activities and therapy exposure

Signature activities were modelled independently in 41 tumour types for each signature. Participants were included if age at sampling, sex and principle components of germline variants were available and the sample was labelled as a primary tumour or a recurrence of a primary tumour (except in the case of melanomas where the sample could be metastatic provided that the primary site was also a melanoma). The 157 chronic lymphoblastic leukaemia participants were excluded. Any male breast, ovarian or uterus tumours were removed as were any female testicular or prostate tumours. The final sample list includes 9,946 tumour samples.

Five covariates were used: log(age), sex (0=male, 1=female) and the first, second and third components of the PCA performed on 55,603 GEL participants (https://cnfl.extge.co.uk/display/GERE/Principal+Components+and+genetically+inferred+relatedness). Covariates were normalised to zero-mean unit variance. Signature activities were modelled using a negative binomial GLM or, where this failed to converge, a poisson GLM (which we’ll refer to as the NB/P model). We also separately used logistic regression model on the binary parameter

$B_{i} = \{0 if A_{i}==0 or A_{i}<median(\{A\}), 1 otherwise\}$

where $A_{i}$ is the signature activity of sample $i$ and the median is estimated over the samples in the tumour type.

The fitted coefficient of the NB/P model is the rate of change of the expected log activity with respect to the normalised covariate

$$\beta= \frac{\partial log(\mu)}{\partial X}$$

where $X$ is the normalised covariate and $\mu$ is the expected mutation rate due to the signature. For the logistic model it’s the rate of change of log-odds of the signature activity being non-zero with respect to the renormalised covariate. Dividing $\beta$ by the square root covariance of the covariate in the group gives the rate of change of log rate with respect to the unnormalised covariate.

*P*-values for the Logistic regression model were evaluated using a Wilks likelihood ratio test. Signature activities are not well represented by negative binomial or poisson distributions such that the association significance was inflated for the NB/P model. The significance of association for the NB/P model was estimated with distilled Conditional Randomisation Testing (dCRT) (Barry *et al.* 2021, Liu *et al.* 2022). The signature activity was first modelled as a function of covariates and the target variable (e.g. treatment exposure) to generate association coefficients. The target variable was also modelled as a function of all other covariates. In the case of treatment exposures this was a binomial model with parameters fit using logistic regression

$${{X'}_{Treatment}}\sim Binom\left( expit\left( Q\alpha\right), n=1 \right)$$

where Q is the design matrix of covariates for all samples of the tumour type where $\alpha$ are the logistic regression fitted parameter values. The target variable values were then resampled from this model 100 times and the z-score for the association with the signature activity was calculated for each resample as

$$Z_{Null} = \frac{dlog(L(X';\beta)}{d\beta} \times\frac{1}{I(X';\beta)}$$

where the likelihood, *L*, is for a NB/P and $I$ is the Fisher information and X’ are the resampled target values. The z-score of the real target variable data is calculated in the same way

$$Z_{Alt} = \frac{dlog(L(X;\beta)}{d\beta} \times\frac{1}{I(X;\beta)}$$

where *X* is the target variable. The *P*-value of association was calculated by comparing the alternative z-score with the distribution of null z-scores using a chi-square test with one degree of freedom based on the mean and variance of the null z-scores.

### Associations between signature activities and DNA repair gene inactivation

Signatures were modelled against DNA repair gene inactivation using the NB/P model with dCRT to evaluate effect sizes and *P*-values as described above. However, the gene inactivation parameter is not binary and must be modelled appropriately. Germline and LoH were resampled from an n=1 binomial GLM

$${{X'}_{Germline}}\sim Binom(expit\left( Q\alpha_{G} \right), n=1), {X'}_{LoH}\sim Binom\left( expit\left( Q\alpha_{LoH} \right), n=1 \right),$$

and somatic hits from a poisson GLM

$${X'}_{Somatic}\sim Poisson\left( exp\left( Q\alpha_{S} \right) \right)$$

where Q is the design matrix of covariates for all samples of the tumour type and $\alpha_{i}$ are the parameters fitted to the logistic models and poisson GLM. The resampled inactivation parameter is the sum of resampled hits with a maximum of 2

$X' = min\left( 2, {X'}_{Germline}+{X'}_{LoH} + {X'}_{Somatic} \right)$.

This was used to generate the null distribution of z-scores as described in the previous section. This method reduced the false positive rate under mock tests (see the next section) to within the expected distribution of *P*-values as shown in **Supplementary Figure 4** where a direct estimation of significance using a Wilks likelihood ratio test would produce a large false positive rate.

### Signature and gene inactivation mock tests

Mock signatures and mock gene inactivations were generated to test for false positives. Three types of mock were generated:

1) Signature activities were simulated for all samples in the dataset from a negative binomial model

$$A_{mock} \sim NB({Q^{T}\alpha}, \theta)$$

where Q is the design matrix of covariates, $\alpha$ are the coefficients and $\theta$ the size parameter of the negative binomial. Covariate coefficients were set to 1 and $\theta=1$ but there was no dependence on the gene inactivation parameter. This model has no hypermutation of samples and can be fitted well by the NB GLM.

2) Gene inactivation values were simulated in three components for the germline, somatic and LoH mutations

$$X_{Germline} \sim Binom(p_{Germline}, n=1), X_{Somatic} \sim Poisson(\mu_{Somatic}), X_{LoH} \sim Binom(p_{LoH}, n=1)$$

where $p_{Germline}, \mu_{Somatic}$ and $p_{LoH}$ were all set to the same value of 0.05, 0.2 and 0.5 for three separate mocks. The gene inactivation parameter was given by

$X = min\left( 2, X_{Germline}+X_{LoH} + X_{Somatic} \right)$.

By simulating the gene inactivations but using the true signature activities, we tested the model on a dataset which is not well represented by a negative binomial or poisson GLM. However, this is akin to the resampling in dCRT which means that the method should not produce false positive inflation by construction.

3) Another set of gene inactivation mocks were generated by perturbation resampling gene inactivations within each tumour type. This was performed for the genes NTHL1, BRCA2 and MGMT which had different mutation rates in different tumour types. These mocks tested whether the method was robust against both hypermutated samples and any variation in the distribution of gene inactivations away from the assumed model used in the dCRT described above.

The three mocks test the method in different ways and can be used to estimate the false positive rate in the results. **Supplementary Fig. 4** shows *P*-*P* plots for the respective mock tests demonstrating the significant reduction in false positive detection achieved using dCRT compared with Wilks likelihood ratio test.

### Associations between signatures and tumour histologies

The status and clinical features of tumours across different cohorts were retrieved from PHE/NCRAS in GEL. TNM stage was used along with NPI for breast cancers, Dukes for CRC, FIGO for ovarian and uterus tumours, Gleeson for prostate and Breslow for skin melanomas. In all cases the grade and stage were treated as continuous variables with stage given values of 0,1,2,3,4 and grade as 1,2,3,4. ER, PR and HER2 hormone status were acquired for breast cancer participants and also binarised such that N=0, P or Pm=1 and any other value is treated as missing.

Associations were performed against the signature activities using logistic regression controlling for age, sex and population principal components of each participant. Missing data reduces the sample size we have to work with. Out of the 9,911 samples with age, sex and principal component information which pass quality control, 7,347 had stage and 6,584 had grade recorded.

Each regression was repeated with and without the clinical feature included as a covariate and Wilks likelihood ratio test generated the *P*-value of association **(Fig.10)**.

### Survival analysis

Survival time for each participant was measured from the data of tumour sampling to the date of most recent follow up or death. We used a Cox proportional hazards model for each tumour type implemented in the python lifelines package with age, sex and principle components as covariates and include tumour grade encoded as a numerical value from 1 to 4. TNM stage was not included as a covariate as this failed proportionality assumptions across multiple cohorts. Signature activities were transformed to $log(activity+1)$ and were also used as covariates when being tested. All variables were normalised to zero mean, unit variance before fitting the model.

In all cases we ran proportional hazard (PH) and Ljung-Box tests and we only report results where the PH-test of the signature coefficient has *P*-value>0.01.
