## Supplementary materials for "Comprehensive repertoire of the chromosomal alteration and mutational signatures across 16 cancer types from 10,983 cancer patients"

#### Artefacts

DBS12 is an artefactual signature caused by T>C mutations in repetitive T regions of the genome where the C is often displaced by a single base due to read misalignment; resulting in clonal T>C mutations being called as heterozygous subclonal TT>CC mutations (reported by Degasperi et al 26 as DBS28; personal communication).

SBS57 is clustered with signatures for dMMR (**Fig. 4**) and is caused by TTT>TCT single base substitutions. This is caused by indels occurring in long homopolymer chains which can trick the variant caller into calling single base substitutions at the end of the homopolymers rather than InDels which occur somewhere in its length.

#### InDel signatures

When we first extracted ID signatures, we used all mutations which passed quality control filters applied by Genomics England. This produced a mutation count matrix of 142,764,499 InDels across all samples. Amongst these mutations was a significant excess of single T/A deletions from TT/AA dinucleotide pairs across all samples (**Supp Fig. 13**). This mutation type is a key feature of ID13, a UV signature which has previously only been extracted in melanomas.

The cause of this excess of mutations are germline single base deletions in long homopolymers which occur adjacent to a TT/AA dinucleotide pair. In the tumour, a single base substitution can occur on one of the bases in the dinucleotide pair. For any single read the exact mutation is degenerate with the scenario where a deletion happens in the dinucleotide pair with a single base substitution at the start of the long homopolymer. The Strelka variant caller typically calls the latter event as the somatic mutation which has taken place as context from germline reads is not taken into account. Therefore a germline single base deletion in a long homopolymer is incorrectly called as a somatic deletion to a homopolymer pair.

We therefore applied an additional filter within Genomics England which removes somatic InDels within 10bp of a germline InDel which is present in >1% of samples in GEL or GNOMAD. This reduced the rate of ID13 in non-melanoma samples by ~99%. By inspecting the reads we found that remaining ID13-attributed deletions were not obviously caused by artefacts in alignment or variant calling.

#### Linearly dependent signatures

As described in the methods, new signatures were added to the COSMIC reference list one at a time if they could not be decomposed to the reference list with $cos(sim)>0.8$. This mimics the approach which was taken to define novel signatures as applied by SigProfilerExtractor. This ensured that any novel signature was linearly independent of all current signatures. However it did not guarantee a set of signatures which were linearly independent of one another. If a signature already included in the reference list could be composed of two novel signatures to be added, we would end up with signatures which are linearly dependent. This occurred with our handling of the SV signatures. SV9 can be composed of a linear sum of SV2, SV4, SV6 and SV13 with $cos(sim)=0.99$ (**Supplementary Table 3**). However, since SV9 was extracted in more tumour cohorts, it was first to be added to the reference list and SV4 and SV6 were added at later iterations.

This issue is present in COSMIC already where signatures added in the past can be composed of signatures which are later added to the list. 51 out of 78 COSMIC SBS signatures can be produced by linear combinations of other COSMIC SBS signatures with $cos(sim)>0.8$. A new approach to producing signature references which can be updated over time is required to allow for robust signature extraction and deconvolution.

#### Mechanistic basis of signatures

In the main text we highlight some key associations found between signatures and DNA repair gene inactivations or treatment exposures in specific tumour groups. One association discussed is the relationship between a clustered group of signatures, SBS17a, SBS17b, SBS18, SBS93, DBS4, DBS7, DBS16, DBS19, ID14 and SV7 with *POLG* inactivation. It has previously been shown that *POLG* is recurrently somatically mutated in colorectal cancer with links to mutagenesis in mitochondrial DNA^1^. However, none of the samples in our cohort have somatic point mutations which were classed as oncogenic by OncoKB and the associations were driven by *POLG* inactivation through LoH.

We assessed the importance of signatures for patient outcome by analysing the relationships between signature activities and tumour stage and grade and overall survival of patients.

The burden of HRD signatures is significantly associated with tumour grade and is inflated in ER and PR-negative breast ductal cancers (**Fig. 6**). This relationship between HRD and hormone receptors has been shown previously when comparing ER+/HER2- cancers with triple-negative breast cancers (TNBC)^1^; however, HER2 status does not show a strong relationship with HRD in our analysis.

HER2-positive breast ductal cancers have higher rates of APOBEC signatures^2,3^ (**Fig. 6b,d**). HER2 promotes cell growth leading to more aggressive tumours^4^ and we find that APOBEC activity is significantly associated with overall patient survival (**Fig. 7b**). This is a novel finding for breast ductal cancers which may have important consequences for patient prognosis and treatment decisions.

MMR signature activity is inflated in colorectal cancers with higher grades but lower stage (**Fig. 6c**). The same relationship is seen with *POLE* signatures in colorectal and uterus cancer. This has been previously observed in gliomas^5^ and other studies have shown that *POLE*-mutated uterus cancers are higher grade^6^ and there has been some previous evidence for higher rates of *POLE* mutations at earlier stages^7^ but this is the first time this effect is reported for colorectal cancer. This is clinically important as *POLE*-mutated uterus cancers show improved progression-free survival^7,8^ however there were not enough *POLE* samples in this work to demonstrate a relationship with patient survival.

#### Confounding of gene-inactivation/treatment associations

Along with the relationships described for gene inactivations and treatment exposures, each signature is typically associated with multiple seemingly unrelated genes in a tumour type although usually with smaller effect sizes. This may be caused by confounding between inactivation of different genes which are highly correlated (**Supp. Fig. 7**) or reverse causation as tumours with higher mutation rates are more likely to have mutations in genes. Many chemotherapeutic agents are administered in combination such that separate treatments are also not independent (**Supp. Fig. 9**). To unambiguously ascribe signature causality requires controlled laboratory experiments or analysis of tumour recurrence^9,10^.

#### Signature-associated survival

In a number of tumour types specific signatures are indicative of patient overall survival. APOBEC activity indicated by SBS2 is associated with reduced survival of chondrosarcoma. APOBEC3B expression has been demonstrated to reduce cell apoptosis in chondrosarcomas leading to positive selection^11^. Our results suggest that SBS2 activity is a powerful indicator for selecting treatments which target APOBEC3B expression.

CN17 is active in 88 out of 296 bladder transitional cell carcinoma (TCC) samples and significantly associated with reduced survival in those patients (P=3.6e-5). Even after controlling for age, sex, population stratification and tumour grade CN17 remains a powerful indicator of patient survival (Cox PH beta=0.49, 95% CI [0.19, 0.80]). CN17 is a signature of HRD and could be important for selecting patients most likely to respond to PARP inhibitor treatment as is being considered for breast ductal patients^12^.

Colorectal cancers with SBS17b activity have significantly worse overall survival (**Fig. 7a,b**). SBS17b is associated with *POLG* inactivation, however, previous work has suggested a relationship with 5-FU treatment^13^. If tumours which require treatment with 5-FU are typically more severe cases then the patients would have worse survival prospects and this would be an example of confounding by indication. The association of DBS5 activity, driven by oxaliplatin exposure, with reduced survival in lung adenocarcinoma and ovarian cancers is likely also caused by this type of confounding.

#### Comparison with Degasperi+2022

Contemporaneously, Degasperi et al. have also analysed 100kGP data reporting the identification of 40 novel SBS and 18 DBS signatures. There are salient differences between our analysis and Degasperi et al. which impact on recovery of signatures. Firstly, we excluded data from 893 PCR-amplified DNA samples to mitigate against sequencing artefacts. Secondly, we excluded 1,470 samples where histology could not be confidently established, or samples had failed sequencing quality control to prevent excess signature extraction resulting from additional variability of mutation rates in tissue types. Thirdly, we did not analyse multiple samples from the same cancer (excluding 531 samples), to guard against overfitting of repeated data. Finally, the signatures reported by Degasperi *et al.* are not linearly independent and require manual interpretation to select input samples, select signature clusters and assign novelty. Such signatures can only be considered for tumour types in which they were extracted since deconvolution methods such as non-negative least squares assume a linearly independent reference.

Using SPE we examined whether the signatures extracted by Degasperi et al could be composed from the COSMIC reference signatures and the novel signatures herein. Of the 40 novel SBS signatures from Degasperi *et al.*, three match the novel SBS signatures we identify; however, 32 of the purported novel signatures can be decomposed to COSMIC signatures (i.e $cos(sim)>0.8$) (**Supp. Fig. 10**). Degasperi *et al.* reported 39 DBS signatures, 30 of which are not in COSMIC. All but one (DBS18) of the remaining signatures can be decomposed to the COSMIC reference or the 8 signatures extracted in this work (**Supp. Fig. 11**).

While COSMIC does not presently catalogue SV signatures, our SV1-SV6 are very similar ($cos(sim)>0.9$) to those reported by Nik-Zainal et al^14^ in WGS analysis of 560 breast cancers. SV7, SV8, SV11 and SV13 match the signatures R7, R10, R8 and R6b reported by Desgasperi et al., 2020 in a WGS study of 3,107 cancers^15^ (**Supp. Fig. 12**).

### Figures

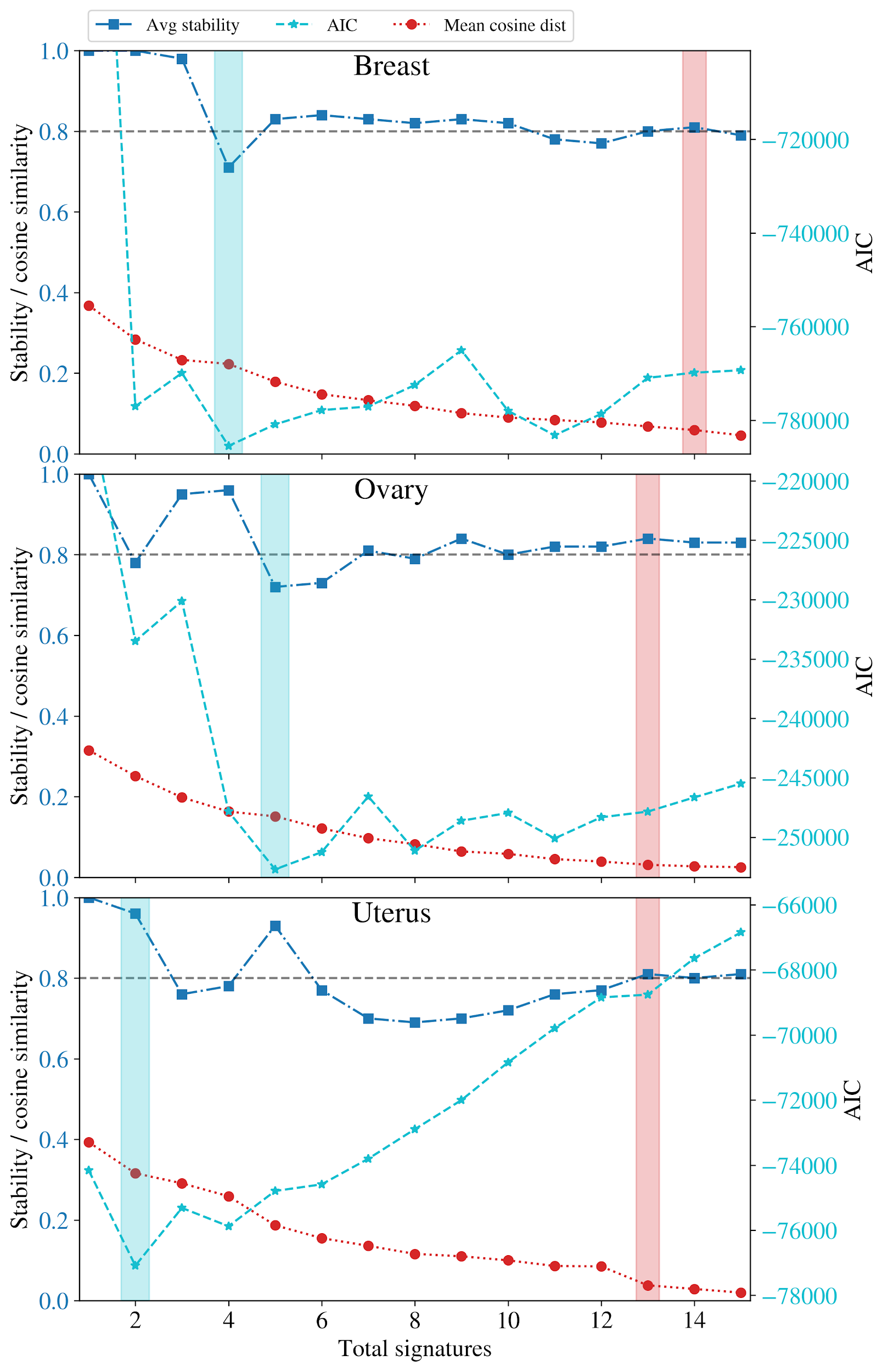

**Supplementary Figure 1.** SigProfilerExtractor picks large numbers of signatures as the optimal solutions for SVs in Breast, Ovarian and Uterus cohorts which include many single-channel signatures. The solution which SigProfilerExtractor picks (red shaded) has average solution stability (blue dot-dashed) greater than 0.8 and looks for significant improvements in signature concordance related to the mean cosine distance (red dotted line). This work uses the AIC (cyan dashed) to select the optimal number of signatures (cyan shaded) in all cohorts. For the three cohorts shown, the AIC solution involves far fewer signatures than the SigProfilerExtractor recommended solution.

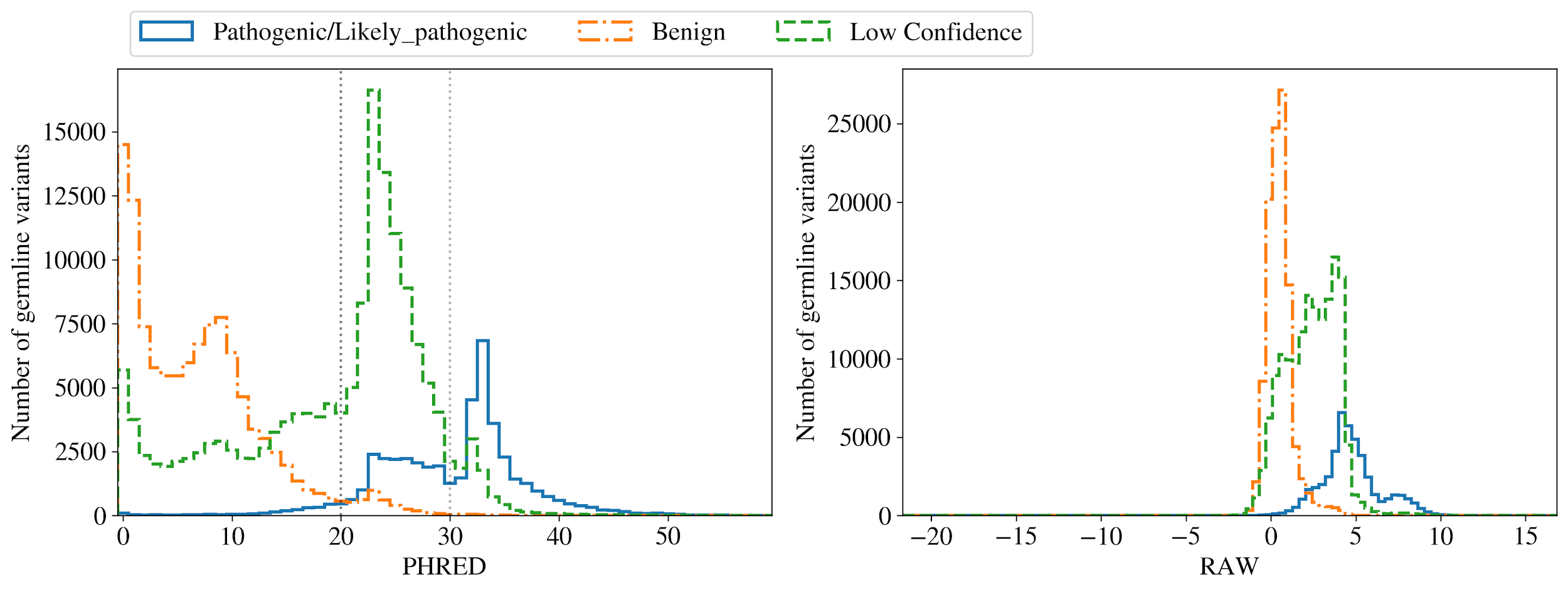

**Supplementary Figure 2.** CADD is run on the set of variants classified in ClinVar producing a set of RAW and PHRED scores. The histograms of these are shown above split by whether ClinVar considers the variant pathogenic, benign or if there is low confidence. A PHRED threshold of 20 removes most benign variants and keeps the majority of pathogenic variants which are typically nonsense or frame-shift. It also keeps a large number of low confidence variants which are mostly missense mutations.

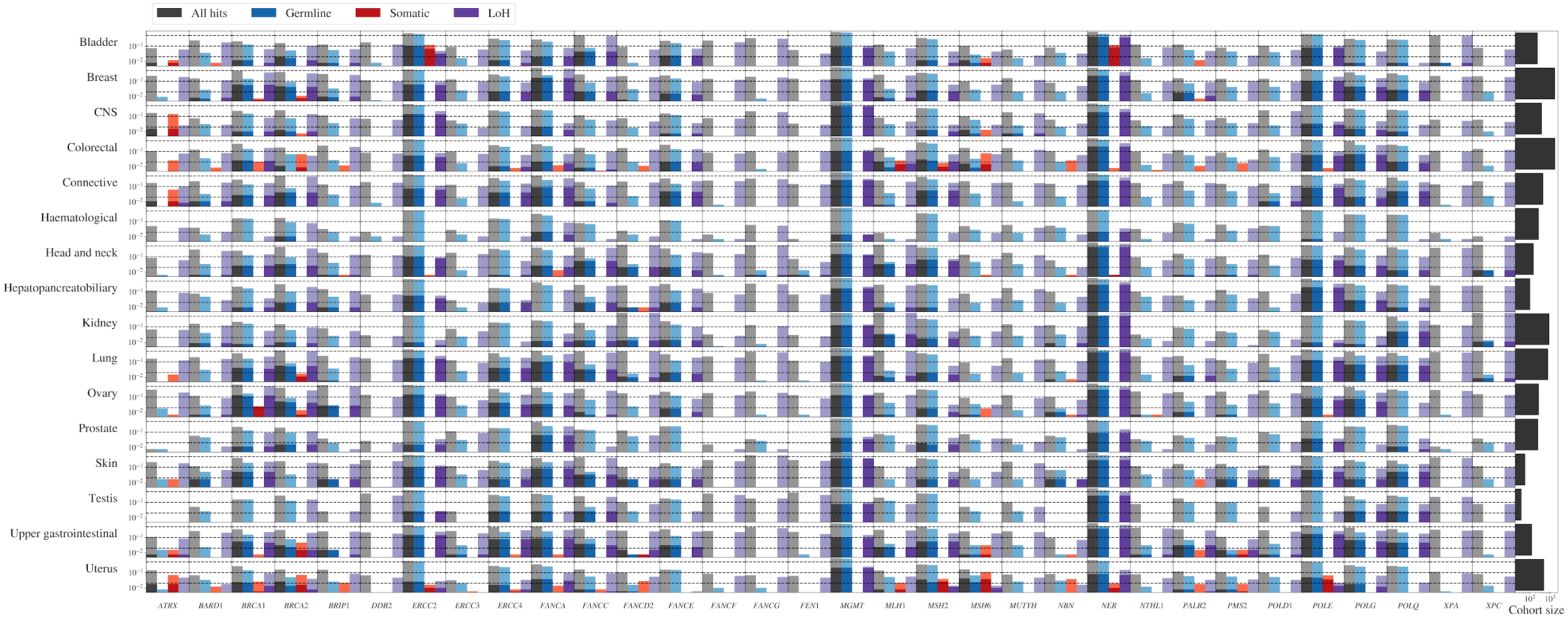

**Supplementary Figure 3.** Gene knockout is determined from a combination of germline, somatic and LoH type mutations. This figure shows the number of samples in each cohort which occur to each gene. The light shaded bars show the number of samples with a single hit in the germline (blue), somatic (red) and LoH (purple). The darker shading for each bar is the subset of samples with one hit of that type which also received a second hit (which may have been of a different type). The grey bars are the totals of single or double hit samples and the histograms on the right show the number of samples in each cohort.

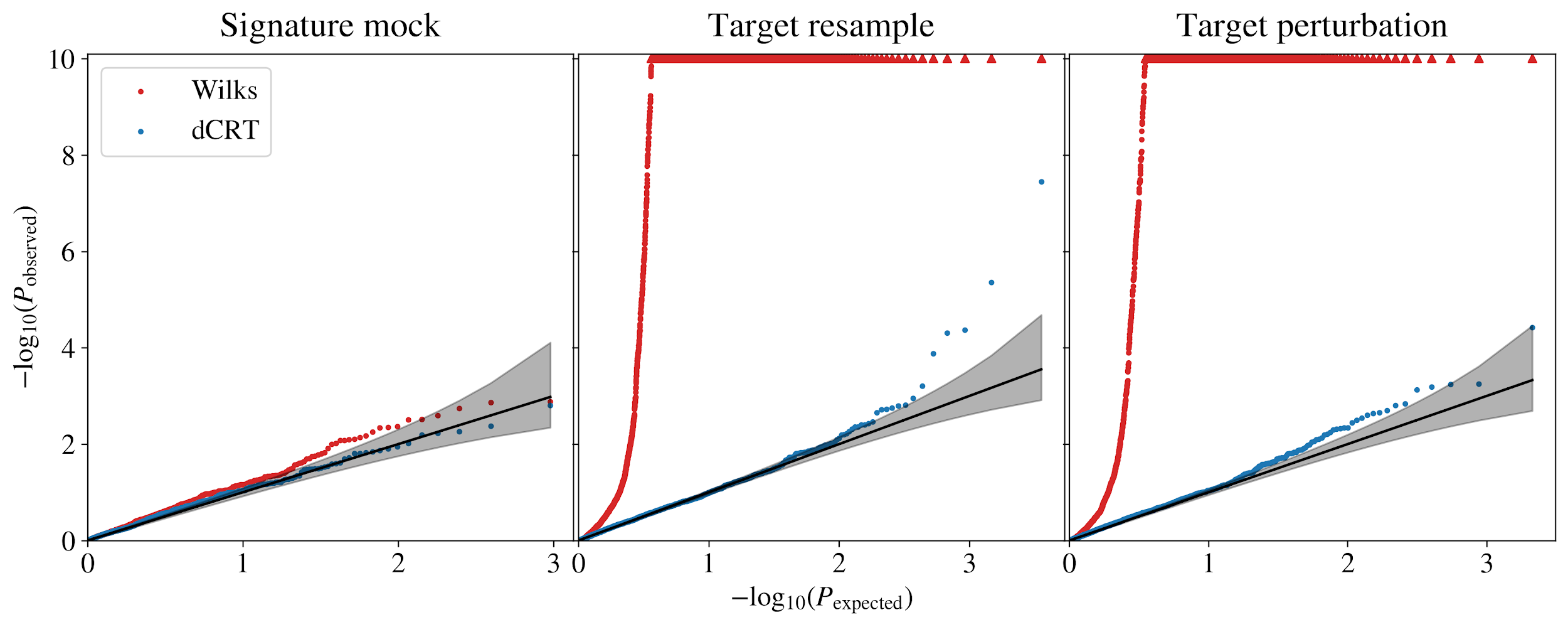

**Supplementary Figure 4.** Mock tests are run for the gene knockout associations using three methods: the signature is resamples from a negative binomial model, the target is resampled from the input target model and the targets undergo a perturbation resampling to reorder (**Methods**). In all cases there should be no association between the targets and signatures. The figures show Q-Q plots of expected and observed *P*-values. When applying a Wilks likelihood ratio test on the negative binomial model outputs (as is done in Liu+2022), there is massive *P*-value inflation resulting in a large Type-I (false positive) error rate. However, conditional resampling successfully reduces this error rate to within the expected uncertainty on the distribution of p-values.

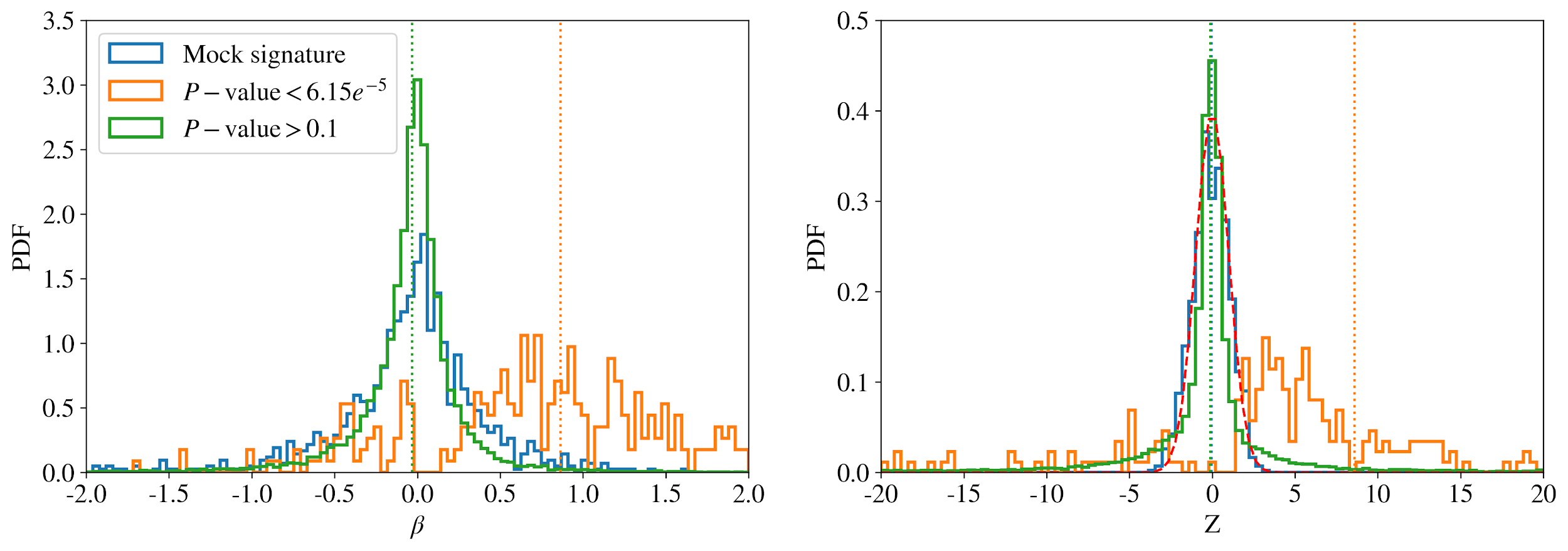

**Supplementary Figure 5.** The distributions of association coefficients and their respective Z-scores are shown for the subset of results with significant *P*-values (orange), for the subset with insignificant *P*-values (green) and the mock samples (blue). For mock and low significance results there is no bias towards a non-zero association coefficient and the z-scores are close to standard normally distributed (red dashed line). The mean of the significant results (orange dotted line) is significantly shifted towards positive values with a mean of Z~8 showing that gene knockout is typically associated with an increase in signature activity as expected.

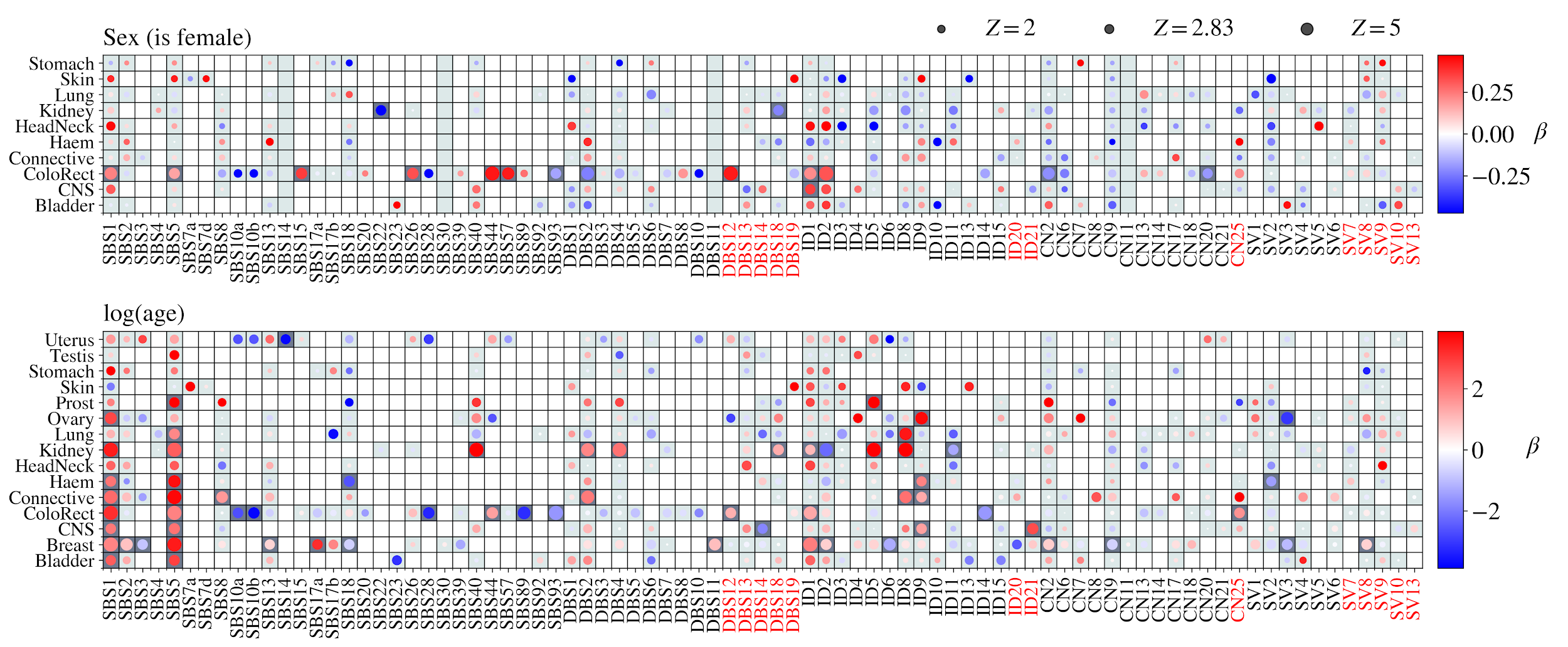

**Supplementary Figure 6. Relationship between signatures with participant sex and age.** Signature activities converted to binary values (presence/absence or above/below median) and modelled against five covariates using logistic regression. The association coefficient (rate of change of log odds) for each covariate, signature and tumour type are shown with the coefficient having been corrected for input parameter normalisation. The fits are only performed for tumour types with at least five samples with non-zero signature activity. Point sizes correspond to z-scores of the log-odds and dark grey grid squares have study-wide significant associations under a chi-square test, which is equivalent to $|Z|>3.42$.

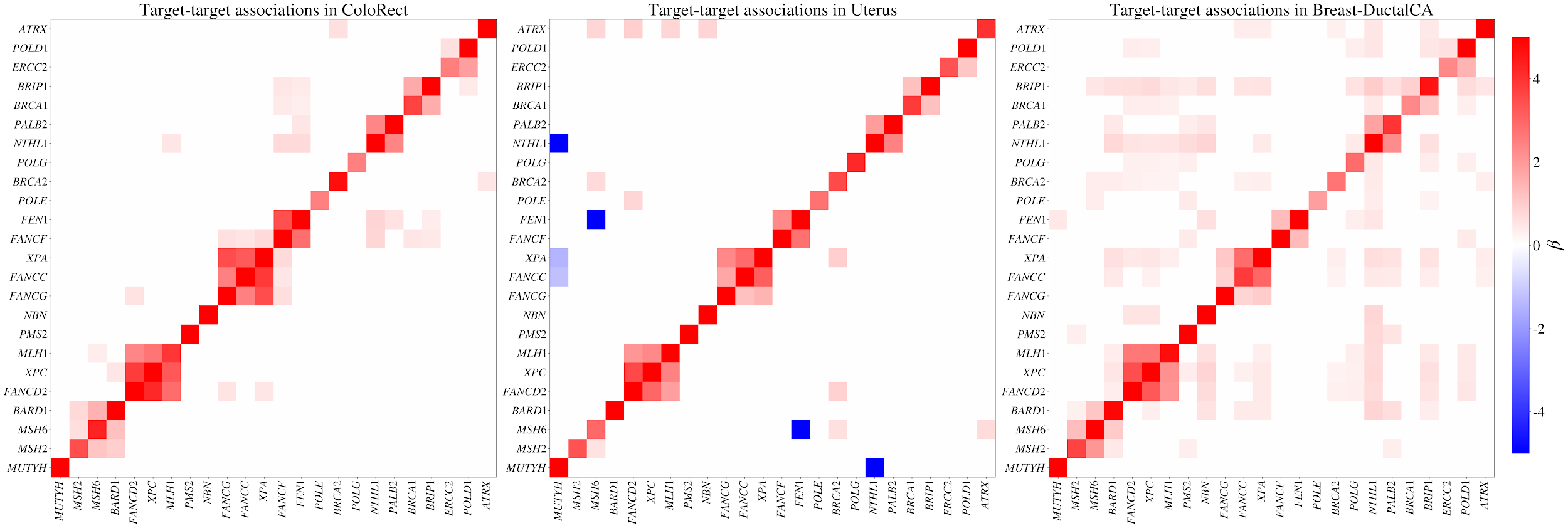

**Supplementary Figure 7.** The knockout of individual genes are not independent but highly correlated in many cases which can complicate interpretation of the results. The genes are associated against one another with a binomial regression with n=2 and the same 5 covariates as are used in the signature regression. The panels show the association coefficients where the chi-squared *P*-value<0.01 in three groups: CRC, Endo and DuctalCA. The genes are ordered by chromosome position and it is clear that nearby genes are often highly associated forming blocks along the diagonal.

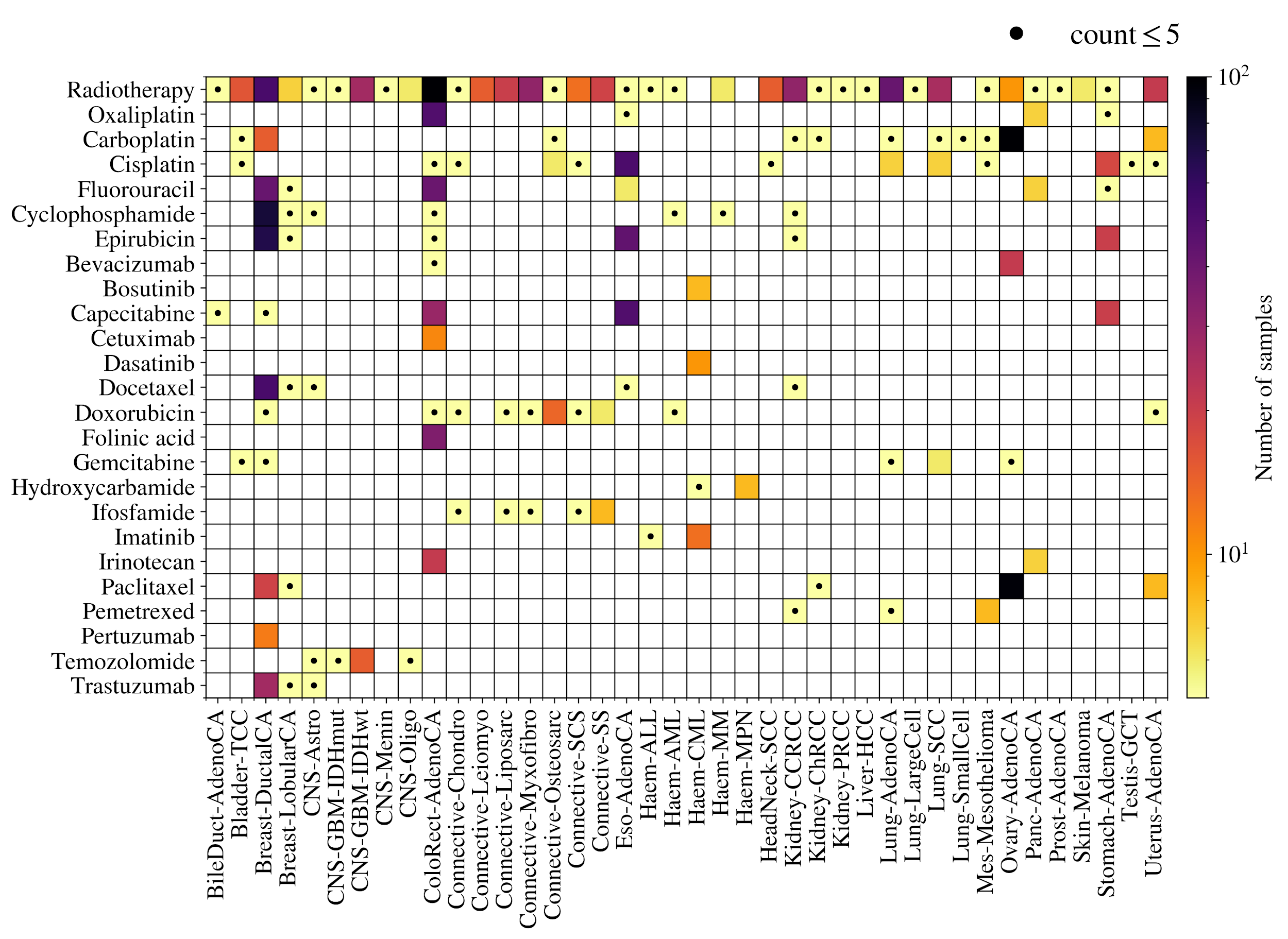

**Supplementary Figure 8.** Information on the treatments used for each tumour are taken from NCRAS and a sample is labelled as exposed to the treatment if the start of the course is before the sample was taken. This figure shows the number of samples exposed to each treatment type in each tumour group where combinations with 5 or fewer samples are labelled with a dot.

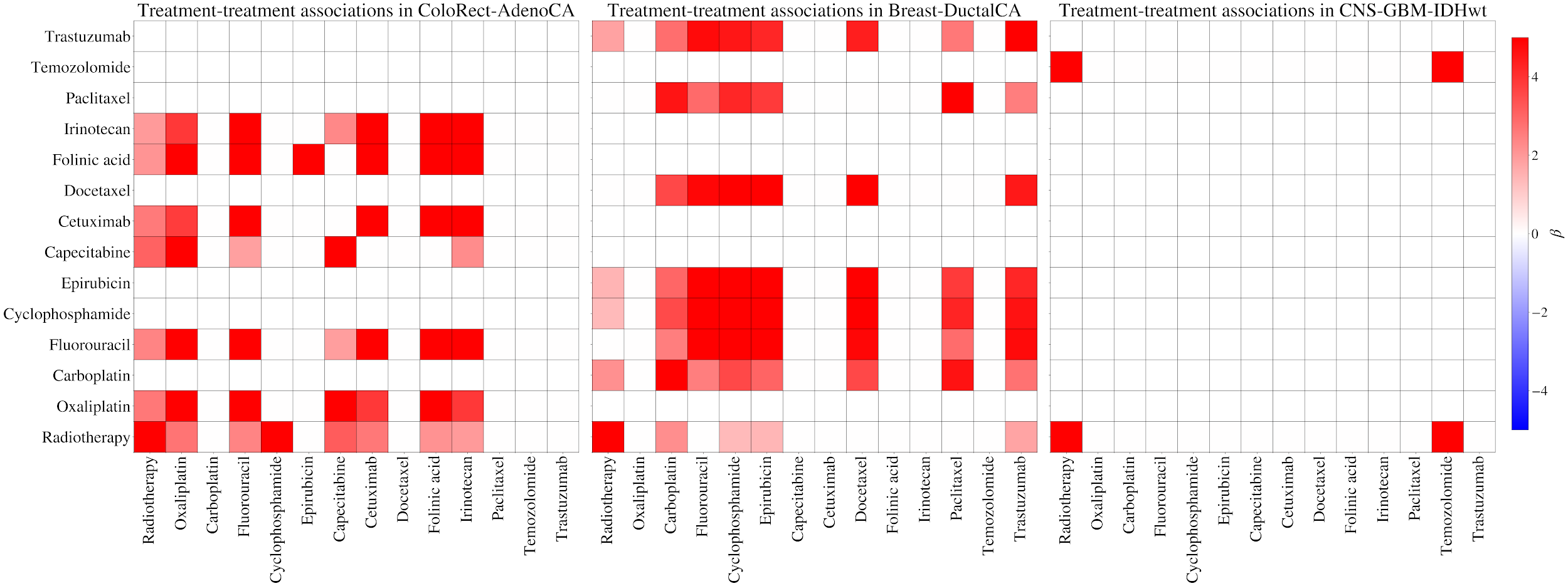

**Supplementary Figure 9.** As in **Supp. Fig. 7**, the associations between treatment exposures are modelled with a logistic regression to find correlations between given treatments in each cohort. CRC, DuctalCA and GBM-IDHwt are shown in the three panels of this figure. Some treatments are typically given together such as epirubicin, cyclophosphamide and fluorouracil in DuctalCA which is given as the combined treatment FEC.

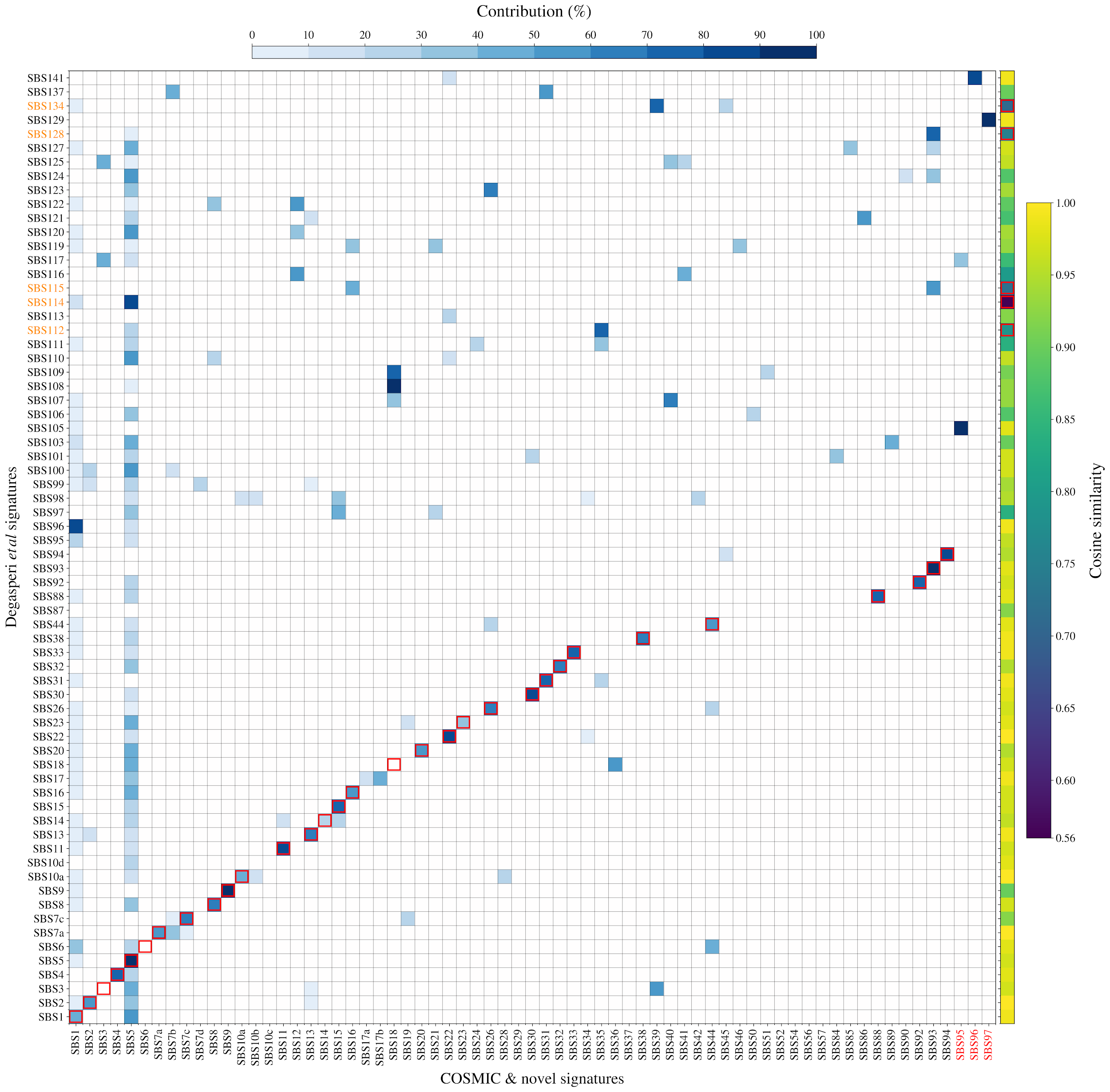

**Supplementary Figure 10**. Degasperi *et al**^16^* SBS ‘green’ rated signatures (high confidence subset) are decomposed to the COSMIC v3 reference signatures and the novel signatures. The blue shading represents the % contribution of each reference signature to the Degasperi signature. The right panel shows that only 5 signatures (highlighted red) could not be decomposed with $cos(sim)>0.8$ between the original signature and the reconstruction. The rest wouldn’t be extracted by SPE as they can be composed of linear combinations of reference signatures.

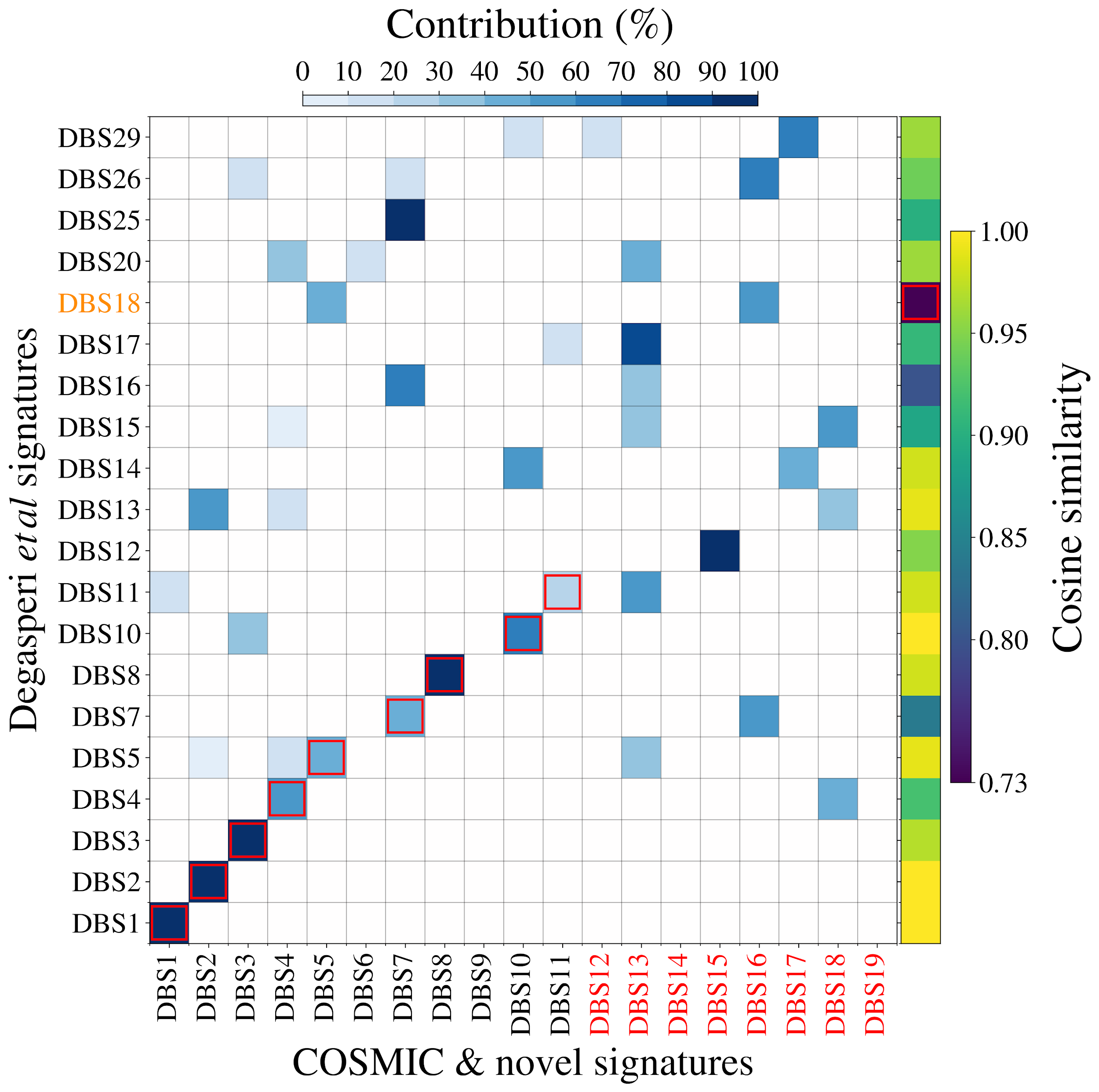

**Supplementary Figure 11**. As **Supp. Fig. 10** but for DBS signatures. Only one Degasperi DBS signature, DBS18, cannot be decomposed to the signatures from COSMIC and this work.

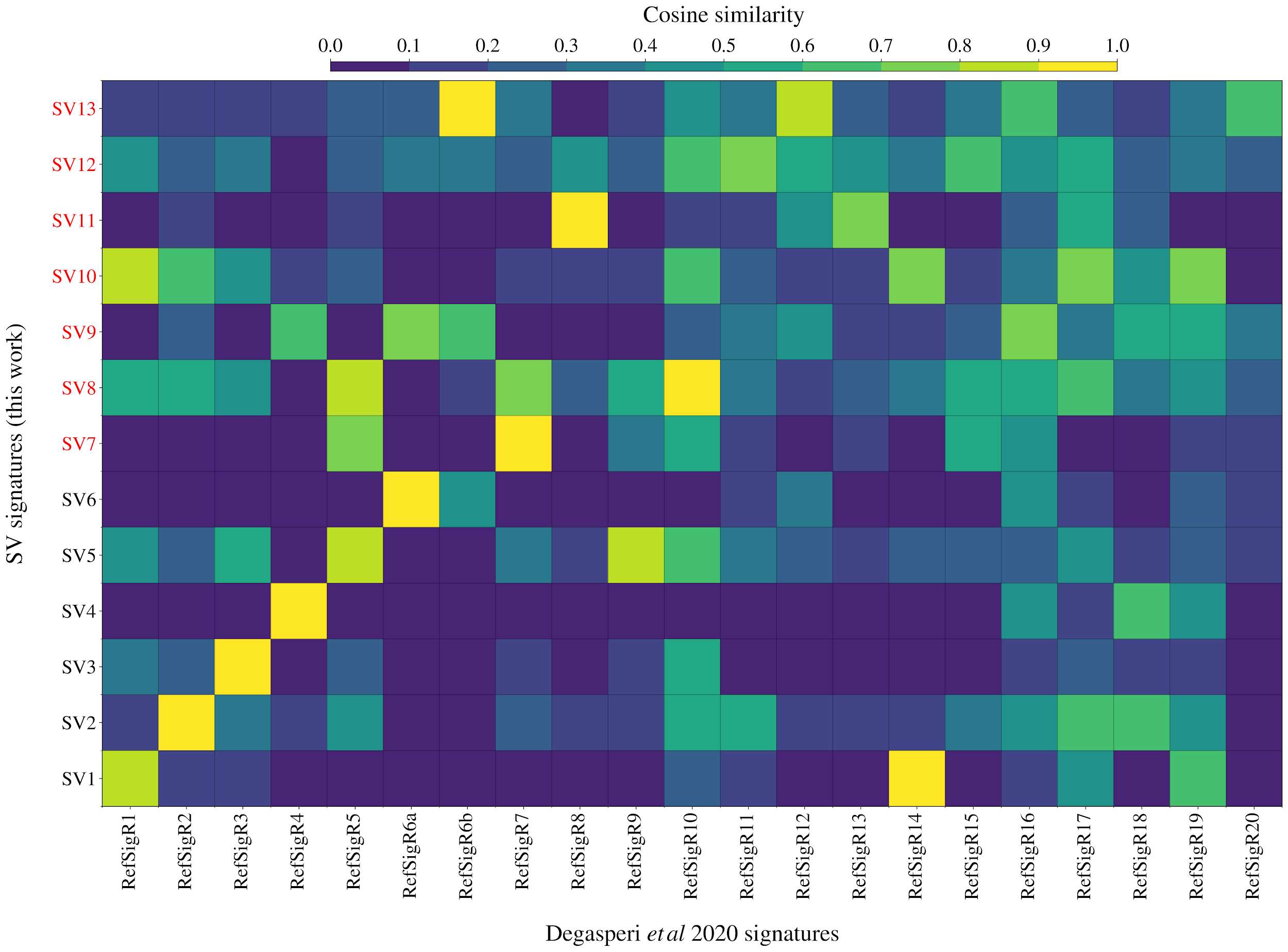

**Supplementary Figure 12.** The 13 extracted SV signatures are matched to previously discovered SV signatures from Degasperi *et al* 2020 based on their cosine similarity. The pairing with the highest similarity is matched where neither signature has already received another match. The red squares show the pairings which are selected where the underlying colour map is the cosine similarity of the pairs.

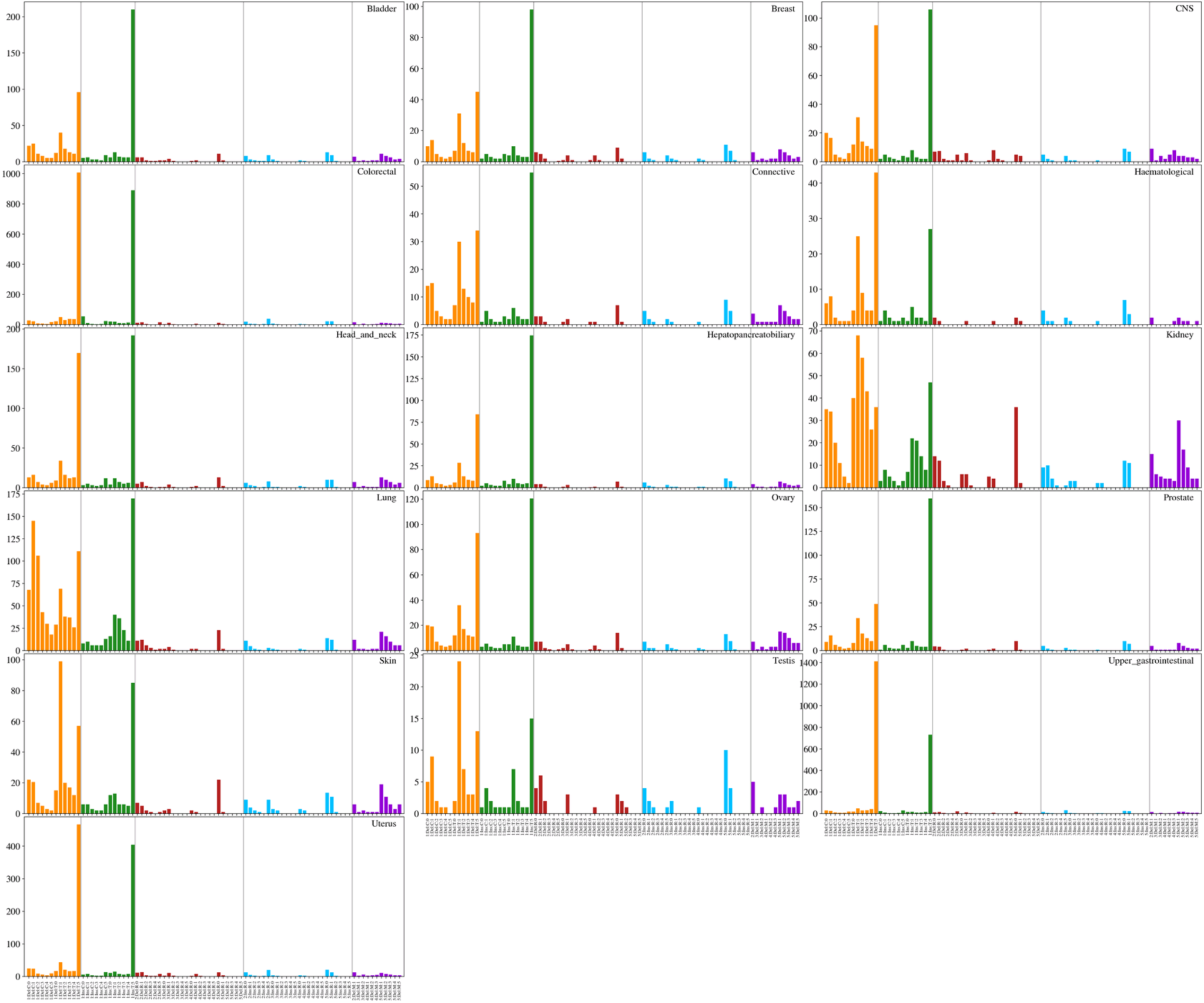

**Supplementary Figure 13.** The median mutation count of each ID mutation type in tumour cohorts used for signature extraction. T deletions in length 2 homopolymers are the largest contribution to skin but are also significant contributors to the majority of other cohorts.
